# Cerebrospinal Fluid Iron Status Predicts Neurocognitive Performance Over Time in Adults with HIV

**DOI:** 10.1101/2020.11.04.20199760

**Authors:** Harpreet Kaur, William S. Bush, Scott L. Letendre, Ronald J. Ellis, Robert K. Heaton, Stephanie M. Patton, James R. Connor, David C. Samuels, Donald R. Franklin, Todd Hulgan, Asha R. Kallianpur

## Abstract

Iron homeostasis is essential for brain health, and iron is also required for HIV replication, but the role of iron is largely unexplored in HIV-associated neurocognitive disorders. These disorders remain extremely common in people with HIV, despite antiretroviral therapy capable of suppressing viral replication, and they involve damage to white matter tracts and synaptodendritic architecture in the brain. We hypothesized that cerebrospinal fluid (CSF) levels of iron and two proteins involved in iron delivery, mitochondrial function, and protection against iron-mediated oxidative stress (H-ferritin and transferrin) are associated with neurocognitive performance over time in adults with HIV. These CSF iron-related biomarkers were measured at baseline (entry) in 403 adults with HIV from a large, prospective observational study who underwent comprehensive neurocognitive assessments at 6-month intervals. All participants were assigned a Global Deficit Score (GDS) and neurocognitive *change* status (*improving/stable/declining*), compared to baseline, at each visit. Biomarker associations with *change* status, and GDS differences over 30, 36, and 42 months of follow-up, were evaluated by multivariable regression. GDS-defined neurocognitive impairment was present in 120 study participants at entry (29.8%); 73% were on antiretroviral therapy. Of 267 participants with longitudinal follow-up, 16% were improving neurocognitively, and 20% were declining at their last follow-up visit (median 36 months). In multivariable-adjusted analyses, higher baseline CSF H-ferritin predicted improving neurocognitive performance at the last assessment (*p*=0.029), and a better GDS over at least 30 months. Higher H-ferritin levels were also associated with better GDS-defined neurocognitive performance over 30, 36, and 42 months in virally suppressed individuals (*p*-values <0.01 at all three time-points) and individuals aged<50 (all *p*-values <0.05). Higher CSF transferrin beneficially influenced GDS-defined neurocognitive performance at all three follow-up visits (all *p*<0.05), particularly in viremic individuals and people with HIV aged 50 and over, but the associations lost statistical significance in analyses restricted to virally suppressed persons. Significant multiplicative interactions with comorbidity were observed for both H-ferritin and transferrin in viremic adults. In summary, higher CSF H-ferritin is associated with better neurocognitive performance over time in adults with HIV, particularly in younger and virally suppressed individuals on antiretroviral therapy. Higher CSF transferrin may be particularly neuroprotective in pro-inflammatory settings such as in older and/or viremic people with HIV. Overall, our findings could reflect better iron delivery to neurons and myelinating oligodendrocytes, better mitochondrial function, and reduced oxidative stress under specific scenarios (*e*.*g*., viremia/aviremia) of HIV infection, and they may provide a rationale for the use of iron-modulating interventions to prevent or treat neurocognitive decline in people with HIV.

## INTRODUCTION

HIV-associated neurocognitive disorder (HAND) remains very common, despite effective combination antiretroviral therapy (ART) [1]. HIV crosses the blood-brain barrier (BBB) early during infection via infected monocytes/macrophages, leading to infection of glia within the brain. While neurons are not infected, sustained glial activation and release of neurotoxic viral proteins promote neuroinflammation and synaptodendritic damage. Risk factors for neurocognitive impairment (NCI) in people with HIV include older age, a lower nadir CD4^+^ T-lymphocyte count, detectable HIV RNA in plasma and/or cerebrospinal fluid (CSF), substance abuse, anemia, and comorbidities [2]. Chronic inflammation due to HIV latency, which is not impacted by ART [1], neuroimmune activation, and the cumulative effects of antiretroviral drugs on mitochondrial function, protein processing, and organellar stress in the central nervous system (CNS) are also implicated [3-6]. Increased risk of NCI associated with the Alzheimer’s disease-related *APOE-ε4* allele has been observed in older adults with HIV on ART [1, 7-9]. Despite intensive research, however, the key pathophysiologic mechanism(s) driving development of NCI when viral replication is suppressed on ART are not well understood.

Iron plays a central role in innate immunity, and its regulation is critical for brain health [10]. The brain has a high metabolic demand for iron, since neuronal development, myelin synthesis and maintenance, and monoamine neurotransmitter synthesis require bioavailable iron; however, iron is also required for HIV replication [10, 11]. While essential for mitochondrial function and diverse metabolic processes, iron can mediate oxidative stress by catalyzing free-radical reactions and must therefore be tightly compartmentalized. H-ferritin, the water-soluble heavy-chain subunit of the iron-storage and transport protein ferritin, crosses the BBB and is the principal source of iron for mature oligodendrocytes [12]. This protein, by virtue of its iron-scavenging properties, also serves an important antioxidant role, preventing iron-mediated oxidative damage [13]. H-ferritin bears high similarity to mitochondrial ferritin, and its reduced expression can lead to oxidative stress, inflammation, and tissue injury [14]. HIV and some antiretroviral drugs dysregulate cellular iron homeostasis, and release of the iron-regulatory hormone hepcidin increases iron sequestration within monocyte-macrophages and glia, benefitting viral growth while limiting its bioavailability to host cells [2]. Functional iron deficiency (reduced bioavailable iron) in the brain, may therefore play an underappreciated role in HIV-associated NCI [15, 16].

This study, in meticulously characterized participants from a large, prospective HIV cohort study, tested the hypotheses that higher levels of CSF biomarkers indicating iron status/iron delivery (H-ferritin and transferrin) are associated with better neurocognitive performance over time, independent of *APOE-ε4*, and that such effects differ by age, comorbidity, and/or viral suppression. Our results suggest that these proteins exert neuroprotective effects in people with HIV under different clinical scenarios (*e*.*g*., viral suppression *versus* ongoing inflammation or viremia), and that iron dysregulation plays a role in HIV-associated NCI in the era of ART.

## RESULTS

### Study Participants

Characteristics of the study population at baseline are presented in **Table 1**. Of 403 individuals evaluated, 120 had Global Deficit Score (GDS)-defined NCI at baseline, and 283 were not impaired. Older age (≥50 years), non-African ancestry, and longer durations of HIV infection, were associated with higher (worse) GDS values (all *p*-values <0.05). Participants with no or minimal comorbidity had significantly lower (better) GDS values (median 0.21 *vs*. 0.42, *p*<0.001). In the entire sample, 294 (72.9%) were on ART at baseline, and 184 (62.5%) of these participants were virally suppressed. Of 398 participants who were genotyped, just under one-third were carriers of the *APOE*-*ε*4 risk allele (see also **Supplementary material, Table S1**), which was not associated with the GDS at baseline.

**Table 1:**
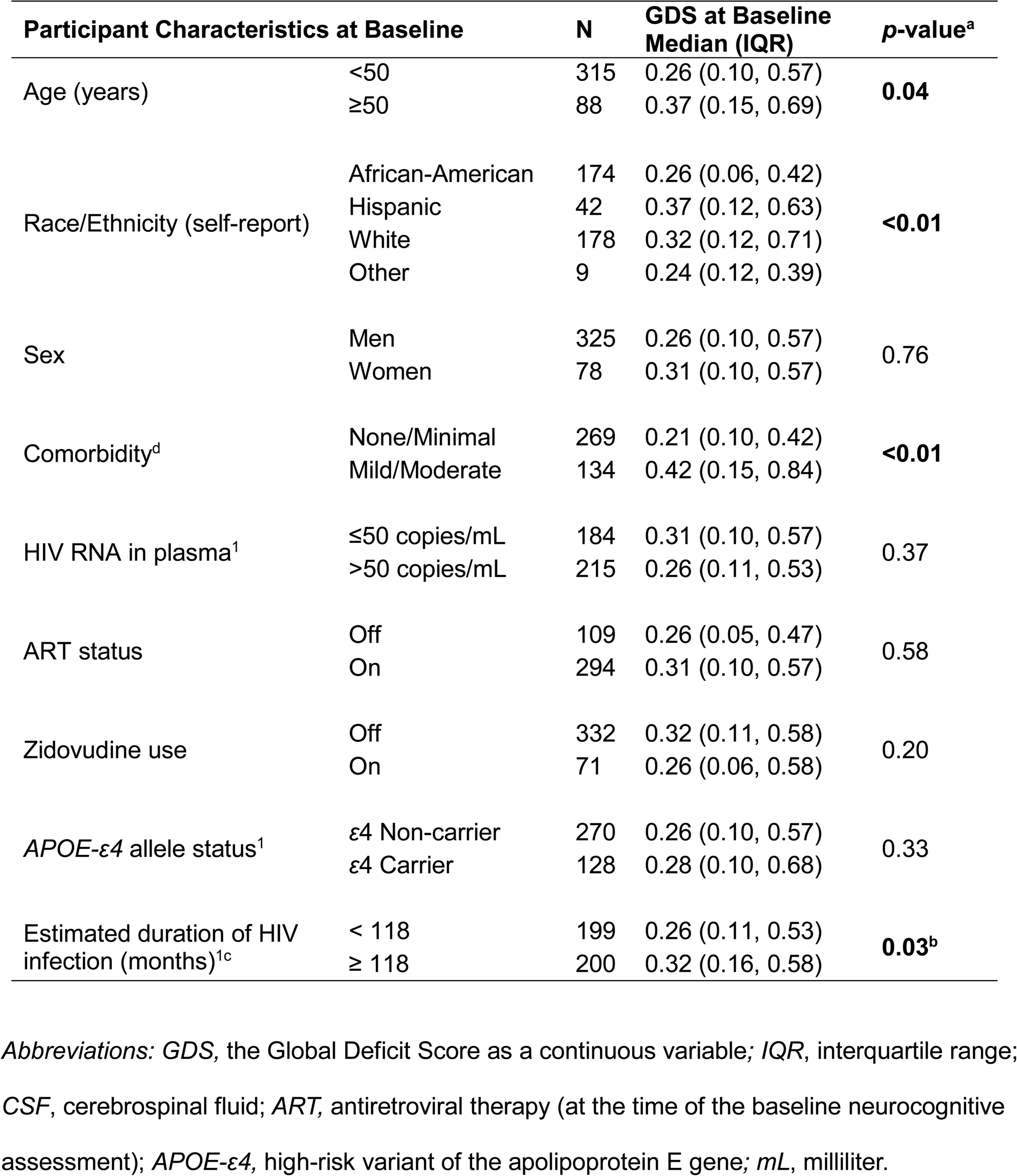
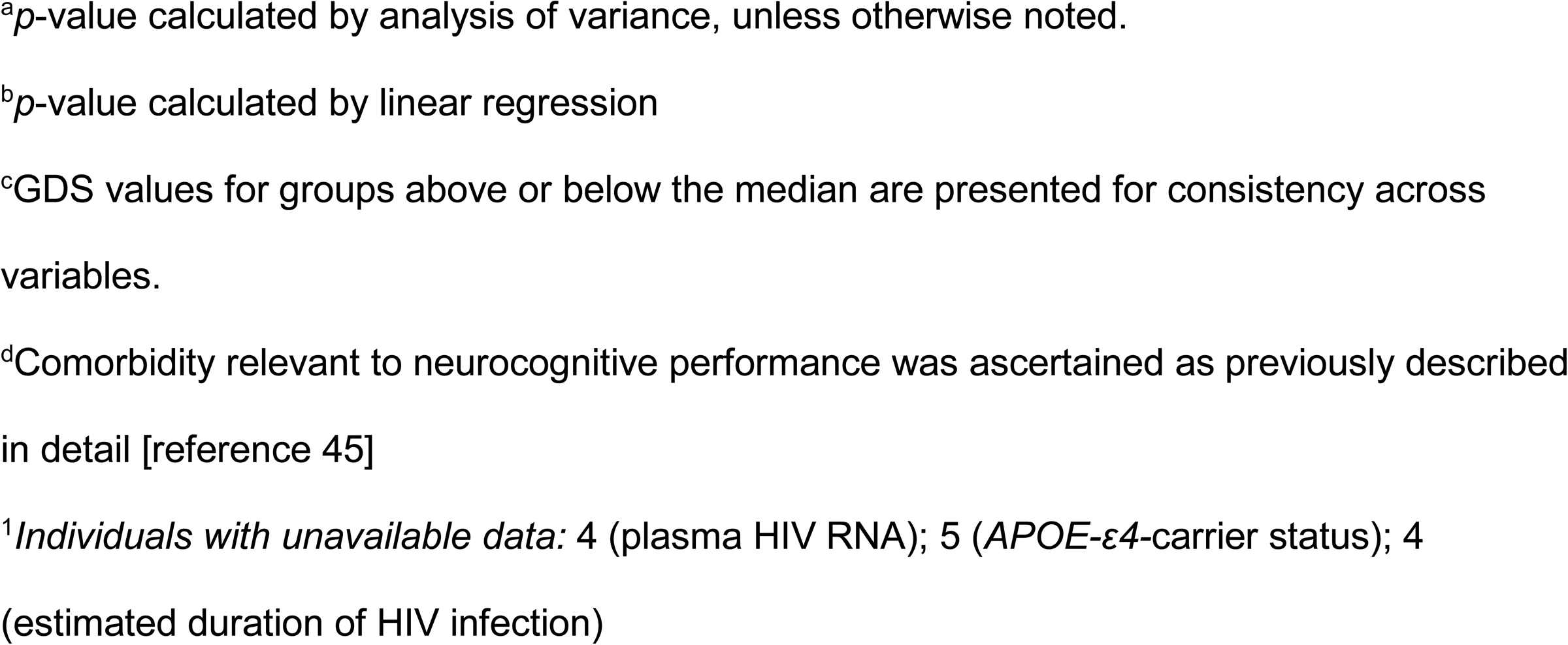
Summary of Global Deficit Score (GDS) values among all CHARTER Study participants at baseline, stratified by demographic and HIV disease characteristics.

| Participant Characteristics at Baseline |  | N | GDS at Baseline Median (IQR) | p-value<sup>a</sup> |
| --- | --- | --- | --- | --- |
| Age (years) | <50 | 315 | 0.26 (0.10, 0.57) | **0.04** |
|  | ≥50 | 88 | 0.37 (0.15, 0.69) |  |
| Race/Ethnicity (self-report) | African-American | 174 | 0.26 (0.06, 0.42) | **<0.01** |
|  | Hispanic | 42 | 0.37 (0.12, 0.63) |  |
|  | White | 178 | 0.32 (0.12, 0.71) |  |
|  | Other | 9 | 0.24 (0.12, 0.39) |  |
| Sex | Men | 325 | 0.26 (0.10, 0.57) | 0.76 |
|  | Women | 78 | 0.31 (0.10, 0.57) |  |
| Comorbidity<sup>d</sup> | None/Minimal | 269 | 0.21 (0.10, 0.42) | **<0.01** |
|  | Mild/Moderate | 134 | 0.42 (0.15, 0.84) |  |
| HIV RNA in plasma<sup>1</sup> | ≤50 copies/mL | 184 | 0.31 (0.10, 0.57) | 0.37 |
|  | >50 copies/mL | 215 | 0.26 (0.11, 0.53) |  |
| ART status | Off | 109 | 0.26 (0.05, 0.47) | 0.58 |
|  | On | 294 | 0.31 (0.10, 0.57) |  |
| Zidovudine use | Off | 332 | 0.32 (0.11, 0.58) | 0.20 |
|  | On | 71 | 0.26 (0.06, 0.58) |  |
| *APOE-ε4* allele status<sup>1</sup> | ε4 Non-carrier | 270 | 0.26 (0.10, 0.57) | 0.33 |
|  | ε4 Carrier | 128 | 0.28 (0.10, 0.68) |  |
| Estimated duration of HIV infection (months)<sup>1c</sup> | < 118 | 199 | 0.26 (0.11, 0.53) | **0.03<sup>b</sup>** |
|  | ≥ 118 | 200 | 0.32 (0.16, 0.58) |  |

*Abbreviations:* GDS, the Global Deficit Score as a continuous variable; IQR, interquartile range; CSF, cerebrospinal fluid; ART, antiretroviral therapy (at the time of the baseline neurocognitive assessment); *APOE-ε4*, high-risk variant of the apolipoprotein E gene; mL, milliliter.
<sup>a</sup>*p*-value calculated by analysis of variance, unless otherwise noted.
<sup>b</sup>*p*-value calculated by linear regression
<sup>c</sup>GDS values for groups above or below the median are presented for consistency across variables.
<sup>d</sup>Comorbidity relevant to neurocognitive performance was ascertained as previously described in detail [reference 45]
<sup>1</sup>*Individuals with unavailable data: 4 (plasma HIV RNA); 5 (APOE-ε4-carrier status); 4 (estimated duration of HIV infection)*

### CSF biomarkers of inflammation and iron status at baseline

Among all 403 study participants, median TNF-α and IL-6 in CSF were 0.44 pg/mL (IQR: 0.33-0.63) and 3.4 pg/mL (IQR: 2.5-4.7), respectively (**Table 2)**. Median CSF iron was 3.1 µg/dL (IQR: 1.6-5.7), H-ferritin was 2.7 ng/mL (IQR: 1.4-4.2), and transferrin was 17.2 (µg/mL, IQR: 10.3-27.6). No significant differences were observed in iron-biomarker levels by comorbidity severity, ART use, or *APOE*-*ε4* genotype, and correlations of CSF iron-related measures with neuro-inflammation (CSF TNF-α and IL-6 levels) and with hemoglobin were weak (all *r*-values <0.20). Levels of both IL-6 and TNF-α were significantly higher in individuals with detectable plasma virus than in virally suppressed individuals, using an HIV RNA cut-off of either <50 copies/mL or <200 copies/mL (all *p-*values <0.01).

**Table 2:** CSF biomarkers of inflammation and iron status in 403 CHARTER study participants at the baseline visit.

| Biomarker | Median (IQR) |
| --- | --- |
| Interleukin-6 (pg/mL)<sup>#</sup> | 3.4 (2.5, 4.7) |
| Tumor-necrosis factor-α (pg/mL) | 0.4 (0.3, 0.6) |
| Iron (µg/dL) | 3.1 (1.6, 5.7) |
| H-Ferritin (ng/mL) | 2.7 (1.4, 4.2) |
| Transferrin (µg/mL) | 17.2 (10.3, 27.6) |

*Abbreviations:* pg/ng/µg/dL/mL, pictograms/ nanograms/ micrograms per deciliter or milliliter; *IQR*, interquartile range; *N*, number.

### CSF iron-biomarker associations with last neurocognitive change status

Among 267 study participants with longitudinal follow-up, median follow-up was 36 months (range 18-54 months). Neurocognitive *change* status at the last assessment was categorized as *improving* in 44 (16%), *stable* in 171 (64%), and *declining* in 52 (20%) compared to the baseline visit. CSF iron and transferrin did not differ by neurocognitive *change* status, either before or after covariate adjustment, whereas people with HIV who were neurocognitively *improving* at last follow-up had higher CSF H-ferritin levels at baseline than individuals who remained *stable*, regardless of impairment status (**Figure 1**). This finding persisted after adjusting for age, plasma HIV RNA, ART, comorbidity severity, and *APOE-ε4* genotype [**Table 3**; Odds Ratio, OR, 1.13 (95% CI, 1.01-1.27); *p*=0.029]. This OR indicates a 13% increase in the likelihood of *improving* neurocognitive performance per unit increase in CSF H-ferritin. Additional adjustment for neuro-inflammation (CSF TNF-α and/or IL-6 at baseline) did not alter these results (see **Supplemental material, Table S2**). CSF iron biomarkers were not significantly different between other groups at the last neurocognitive assessment. It is important to note, however, that all three *change* status groups included both neurocognitively impaired and unimpaired persons.

**Table 3:** Distributions of baseline CSF iron, H-ferritin, and transferrin among all neurocognitively *improving*, *stable* and *declining* individuals at their last neurocognitive assessment visit (median follow-up 36 months, range 18-54 months).

| All Study Participants with Longitudinal Follow-up (N=267) |  |  |  |  |  |  |  |  |  |
| --- | --- | --- | --- | --- | --- | --- | --- | --- | --- |
| Biomarker | Median level (IQR) |  |  | OR (95% CI) |  | OR (95% CI) |  | OR (95% CI) |  |
|  | *Improving*<br>(N=44) | *Stable*<br>(N=171) | *Declining*<br>(N=52) | *Improving vs.*<br>*Stable* | *p* | *Declining vs.*<br>*Stable* | *p* | *Improving vs.*<br>*Declining* | *p* |
| Iron | 3.9<br>(1.9, 7.0) | 3.3<br>(1.4, 6.0) | 2.8<br>(1.8, 5.4) | 1.02 (0.97 - 1.08) | 0.46 | 0.97 (0.89 - 1.05) | 0.39 | 1.07 (0.96 - 1.18) | 0.19 |
| H-Ferritin | 3.0<br>(2.0, 5.4) | 2.7<br>(1.3, 4.2) | 2.7<br>(1.7, 3.8) | 1.13 (1.01 - 1.26) | **0.03** | 1.04 (0.92 - 1.17) | 0.57 | 1.09 (0.95 - 1.25) | 0.20 |
| Transferrin | 17.8<br>(9.0, 29.5) | 16.5<br>(9.2, 28.6) | 17.8<br>(11.8, 27.1) | 1.00 (0.97 - 1.02) | 0.81 | 1.01 (0.99 - 1.03) | 0.16 | 0.98 (0.95 - 1.01) | 0.26 |

**Note:** The *stable* group includes both neurocognitively impaired and unimpaired persons.
Units of measurement: CSF iron (µg/dL), H-ferritin (µg/mL) and transferrin (ng/mL).
*p*-values<0.05 (**bolded**) are statistically significant.
*p*-values were adjusted for age, comorbidity severity, plasma HIV RNA concentration, *APOE-ε4* carrier status (for analyses of CSF iron and H-ferritin) and zidovudine use (for analyses of CSF transferrin).
*Abbreviations:* OR, Odds Ratio; CI, Confidence Interval; IQR, Interquartile range; *p*, *p*-value

**Figure 1.**
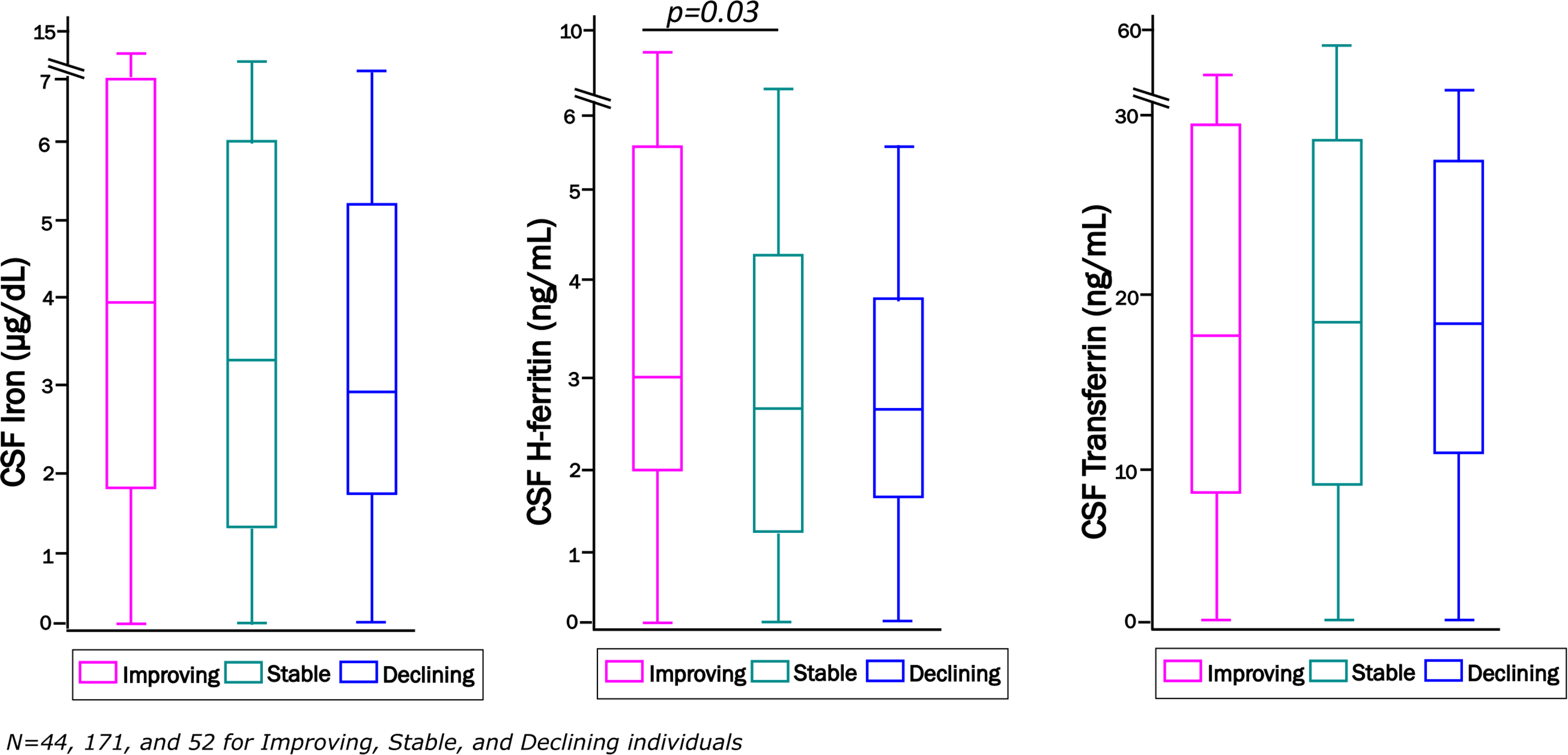
Distribution of CSF iron biomarkers in CHARTER study participants who were defined as neurocognitively *improving*: N=44 (pink), *stable*: N=171 (green) or *declining*: N=52 (blue) at their last follow-up visit. Units of measurements are: CSF iron (µg/dL), H-ferritin (ng/mL), and transferrin (µg/mL). * Statistically significant difference, *p*<0.05.

People with HIV aged<50 years with higher CSF H-ferritin at baseline were 17% more likely to be *improving* at the last follow-up visit per unit increase in this biomarker than neurocognitively *stable* individuals (adjusted OR 1.17, 95% CI 1.03-1.33; *p*=0.016, **Supplementary material, Table S3**). Interestingly, there were no statistically significant differences in H-ferritin by last neurocognitive *change* status in older participants. CSF transferrin and iron did not differ significantly by last neurocognitive *change* status in either age group. *(See also longitudinal age-stratified analysis results, discussed below*.)

### Multivariable-adjusted associations of CSF iron biomarkers with GDS differences over time

Stable changes in neurocognitive performance may require longer periods of time to manifest. For this analysis, we therefore identified 157 individuals with complete longitudinal assessment data at 30 months, 131 of whom had follow-up at 36 months; 110 of the same individuals were also assessed at 42 months. Changes in the GDS over 30-42 months of follow-up for study participants in the highest and lowest CSF H-ferritin and transferrin tertiles at baseline are plotted in **Figures 2** and **3**, respectively, for ease of interpreting directions of effects. CSF H-ferritin tertile had a statistically significant impact on the GDS in all people with HIV: levels in the highest tertile (≥3.55 ng/mL) were associated with lower (better) GDS values over 30 and 42 months of follow-up than levels in the lowest tertile (≤2.15 ng/mL; **Figure 2**, panel ***A***) and **Table 4** (panel ***A***); uncorrected *p-*values 0.037 at 30 months and 0.043 at 42 months for H-ferritin; Huynh-Feldt-corrected *p*-values 0.049 and 0.084, respectively). The absolute magnitude of these effects was small, with *eta*-squared values suggesting that approximately 1 percent of the variance in the GDS over 42 months was explained by H-ferritin tertile at baseline. The H-ferritin association was particularly strong among individuals with no or minimal comorbidity, in whom CSF H-ferritin in the highest tertile was associated with significantly better neurocognitive performance (lower GDS values) over 42 months, compared to participants in the lowest tertile (**Figure 2**, panel ***B***; Huynh-Feldt-corrected *p-*values 0.024, 0.032 and 0.020 at 30, 36 and 42 months, respectively). Although H-ferritin levels did not show the same association in the subset with more clinically significant comorbidity (*data not presented*), an interaction term with comorbidity was not statistically significant in this model. Importantly, in analyses confined to virally suppressed study participants (plasma HIV RNA <200 copies/mL), CSF H-ferritin levels were also associated with significant differences in neurocognitive performance (**Figure 2**, panel ***C***, and **Table 4**, panel ***B***). Aviremic individuals in the highest tertile had significantly better GDS values than individuals in the lowest tertile over 30, 36, and 42 months of follow-up (uncorrected and Huynh-Feldt-corrected *p-*values <0.01 at all three time-points).

**Table 4A.** Associations of CSF H-ferritin levels with GDS differences in all study participants who had comprehensive follow-up for at least 30 months.

| Variable | 30 months (N=75) |  | 36 months (N=62) |  | 42 months (N=48) |  |
| --- | --- | --- | --- | --- | --- | --- |
| | <sup>#2</sup>$p$ | <sup>\$2</sup>$p$ | <sup>#2</sup>$p$ | <sup>\$2</sup>$p$ | <sup>#2</sup>$p$ | <sup>\$2</sup>$p$ |
| H-Ferritin | **0.04** | **0.05** | 0.11 | 0.16 | **0.04** | 0.08 |
| Comorbidity | 0.43 | 0.42 | 0.56 | 0.51 | 0.49 | 0.45 |
| H-Ft*Comorbidity Interaction | 0.26 | 0.28 | 0.29 | 0.31 | 0.13 | 0.17 |
| Plasma HIV RNA | 0.94 | --- | 0.91 | --- | 0.84 | --- |

$p$ -values<0.05 (**bolded**) are statistically significant.
Additional adjustment for *APOE-ε4* carrier status at 30, 36, and 42 months, respectively, did not alter results ( $p$ -values for H-ferritin=0.02, 0.04, 0.03), TNF-α ( $p$ -values for H-ferritin=0.04, 0.12, 0.04) or IL-6 levels ( $p$ -values for H-ferritin=0.04, 0.12, 0.04).
<sup>#1</sup> $p$ -values were adjusted for comorbidity severity, plasma HIV RNA, and an H-ferritin (H-Ft)\*comorbidity interaction term.
<sup>\$</sup> $p$ -values incorporate the Huynh-Feldt correction.

**Table 4B.** Associations of CSF H-ferritin levels with GDS differences in the virally suppressed subset (plasma HIV RNA <200 copies/mL).

| Variable | 30 months (N=75) |  | 36 months (N=62) |  | 42 months (N=48) |  |
| --- | --- | --- | --- | --- | --- | --- |
| | <sup>#2</sup>*p* | <sup>\$2</sup>*p* | <sup>#2</sup>*p* | <sup>\$2</sup>*p* | <sup>#2</sup>*p* | <sup>\$2</sup>*p* |
| H-Ferritin | **<0.001** | **<0.01** | **<0.001** | **<0.001** | **<0.001** | **<0.01** |
| Comorbidity | 0.37 | 0.36 | 0.36 | 0.35 | 0.16 | 0.20 |
| H-Ft*Comorbidity Interaction | 0.33 | 0.33 | 0.34 | 0.35 | 0.44 | 0.42 |

Additional adjustment for either baseline CSF TNF-α (*p*-value for H-ferritin <0.01) or IL-6 (*p*-value for H-ferritin <0.05) did not significantly alter results at 30, 36, and 42 months, respectively *p*-values<0.05 (**bolded**) are statistically significant.
<sup>#2</sup>*p*-values were adjusted for comorbidity and an H-ferritin (H-Ft)\*comorbidity interaction term.
<sup>\$</sup>*p*-values incorporate the Huynh-Feldt correction.

**Figure 2.**
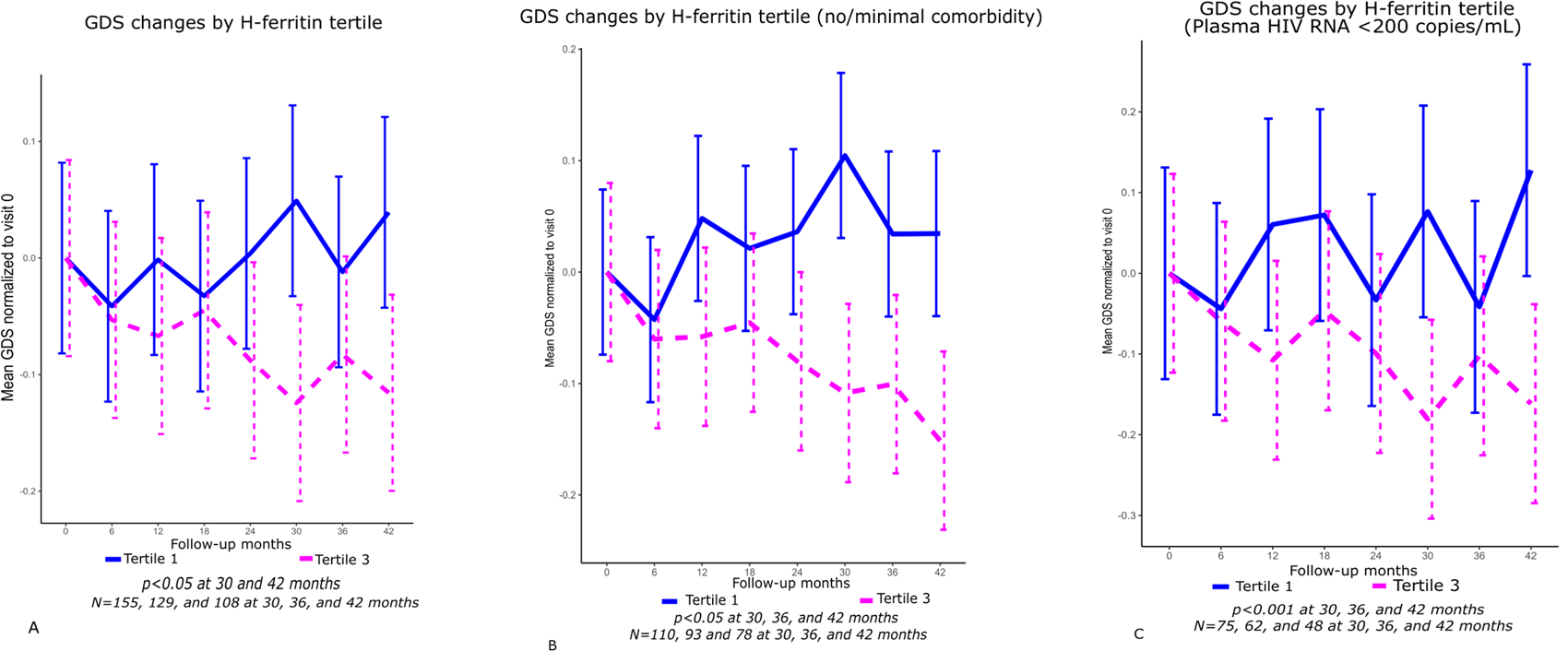
Changes in the Global Deficit Score (GDS) over time in tertiles 3 *vs*. 1 of CSF H-ferritin, in all study participants (**panel *A***, *p*<0.05 for H-ferritin at 30 and 42 months, adjusting for comorbidity and plasma HIV RNA). **Panel *B*** shows changes in the GDS in participants with no comorbidity or minimal comorbidity (*p*<0.05 for H-ferritin at 30, 36 and 42 months), adjusting for plasma HIV RNA. **Panel *C*** shows changes in the GDS by H-ferritin tertile among virologically suppressed participants (*p*<0.01 at 30, 36 and 42 months adjusting for comorbidity).Tertile 2 is omitted for clarity from these plots but all tertiles were included in ANOVA analyses. The mean GDS at each visit was normalized to the baseline GDS for individuals in each tertile. (Results were unchanged even after repeating the analysis, excluding tertile 2, shown in **Supplemental Material, Figure S1**.)

CSF transferrin levels at baseline also had a significant impact on neurocognitive performance over 30-42 months of follow-up, as shown in **Figure 3**. Higher levels (tertile 3, ≥24.0 µg/mL *vs*. tertile 1, ≤12.5 µg/mL) were associated with better neurocognitive function in all study participants over 30, 36, and 42 months (both uncorrected and corrected *p-*values <0.01; **Figure 3** and **Table 5** (panel ***A***). A statistically significant, multiplicative transferrin-by-comorbidity interaction was also observed at all visit time-points (all *p*<0.05). In contrast to H-ferritin, the highest CSF transferrin tertile was associated with better (lower) GDS values only in individuals who had mild or moderate comorbidity. Furthermore, higher CSF transferrin was strongly associated with better GDS values in the subset with detectable HIV RNA in plasma (≥200 copies/mL), as shown in **Table 5**, panel ***B*** (all uncorrected and corrected *p-*values <0.001 for associations with GDS differences over 30, 36, and 42 months). Again, highly significant transferrin-by-comorbidity interaction effects were observed at all three time-points in viremic individuals. This biomarker was not associated with GDS in virally suppressed persons.

**Table 5A.**
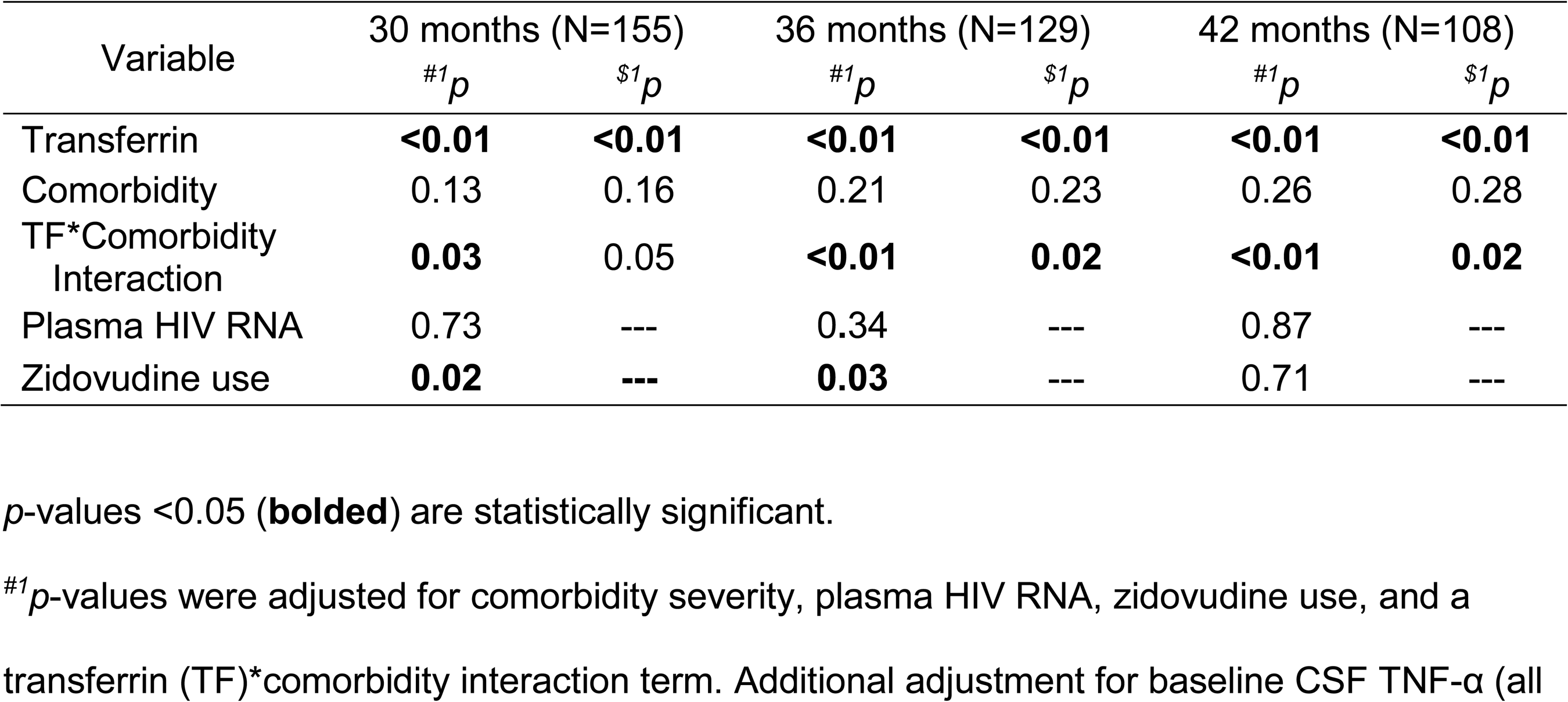
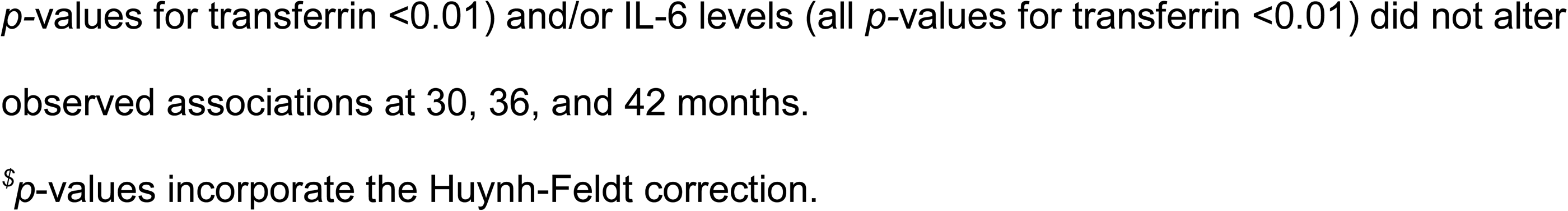
Associations of CSF transferrin with GDS differences in all study participants across all three evaluated visits (up to 30, 36, or 42 months).

| Variable | 30 months (N=155) |  | 36 months (N=129) |  | 42 months (N=108) |  |
| --- | --- | --- | --- | --- | --- | --- |
| | <sup>#1</sup>$p$ | <sup>\$1</sup>$p$ | <sup>#1</sup>$p$ | <sup>\$1</sup>$p$ | <sup>#1</sup>$p$ | <sup>\$1</sup>$p$ |
| Transferrin | **<0.01** | **<0.01** | **<0.01** | **<0.01** | **<0.01** | **<0.01** |
| Comorbidity | 0.13 | 0.16 | 0.21 | 0.23 | 0.26 | 0.28 |
| TF*Comorbidity Interaction | **0.03** | 0.05 | **<0.01** | **0.02** | **<0.01** | **0.02** |
| Plasma HIV RNA | 0.73 | --- | 0.34 | --- | 0.87 | --- |
| Zidovudine use | **0.02** | --- | **0.03** | --- | 0.71 | --- |

$p$ -values $< 0.05$ (**bolded**) are statistically significant.
<sup>#1</sup> $p$ -values were adjusted for comorbidity severity, plasma HIV RNA, zidovudine use, and a transferrin (TF)\*comorbidity interaction term. Additional adjustment for baseline CSF TNF-α (all
$p$ -values for transferrin $<0.01$ ) and/or IL-6 levels (all $p$ -values for transferrin $<0.01$ ) did not alter observed associations at 30, 36, and 42 months.
$^{\$}p$ -values incorporate the Huynh-Feldt correction.

**Table 5B.** Associations of CSF transferrin with GDS differences in the subset with viremia (plasma HIV RNA ≥200 copies/mL).

| Variable | 30 months (N=80) |  | 36 months (N=67) |  | 42 months (N=60) |  |
| --- | --- | --- | --- | --- | --- | --- |
| | $^{\#2}p$ | $^{\$2}p$ | $^{\#2}p$ | $^{\$2}p$ | $^{\#2}p$ | $^{\$2}p$ |
| Transferrin | **$<0.0001$** | **$<0.001$** | **$<0.0001$** | **$<0.0001$** | **$<0.0001$** | **$<0.0001$** |
| Comorbidity | **$<0.001$** | **$<0.001$** | **$<0.0001$** | **$<0.0001$** | **$<0.001$** | **$<0.0001$** |
| TF*Comorbidity Interaction | **$<0.001$** | **$<0.001$** | **$<0.0001$** | **$<0.001$** | **$<0.001$** | **$<0.001$** |
| Zidovudine use | 0.94 | --- | 0.95 | --- | 0.97 | --- |

$p$ -values $<0.05$ (**bolded**) are statistically significant.
$^{\#2}p$ -values were adjusted for comorbidity severity, zidovudine use, and a transferrin (TF)\*comorbidity interaction term. Again, additional adjustment for baseline CSF TNF-α ( $p$ -values for transferrin $<0.01$ ) or IL-6 levels ( $p$ -values $<0.01$ ) did not alter transferrin associations at 30, 36, and 42 months.
$^{\$}p$ -values incorporate the Huynh-Feldt correction.

**Figure 3.**
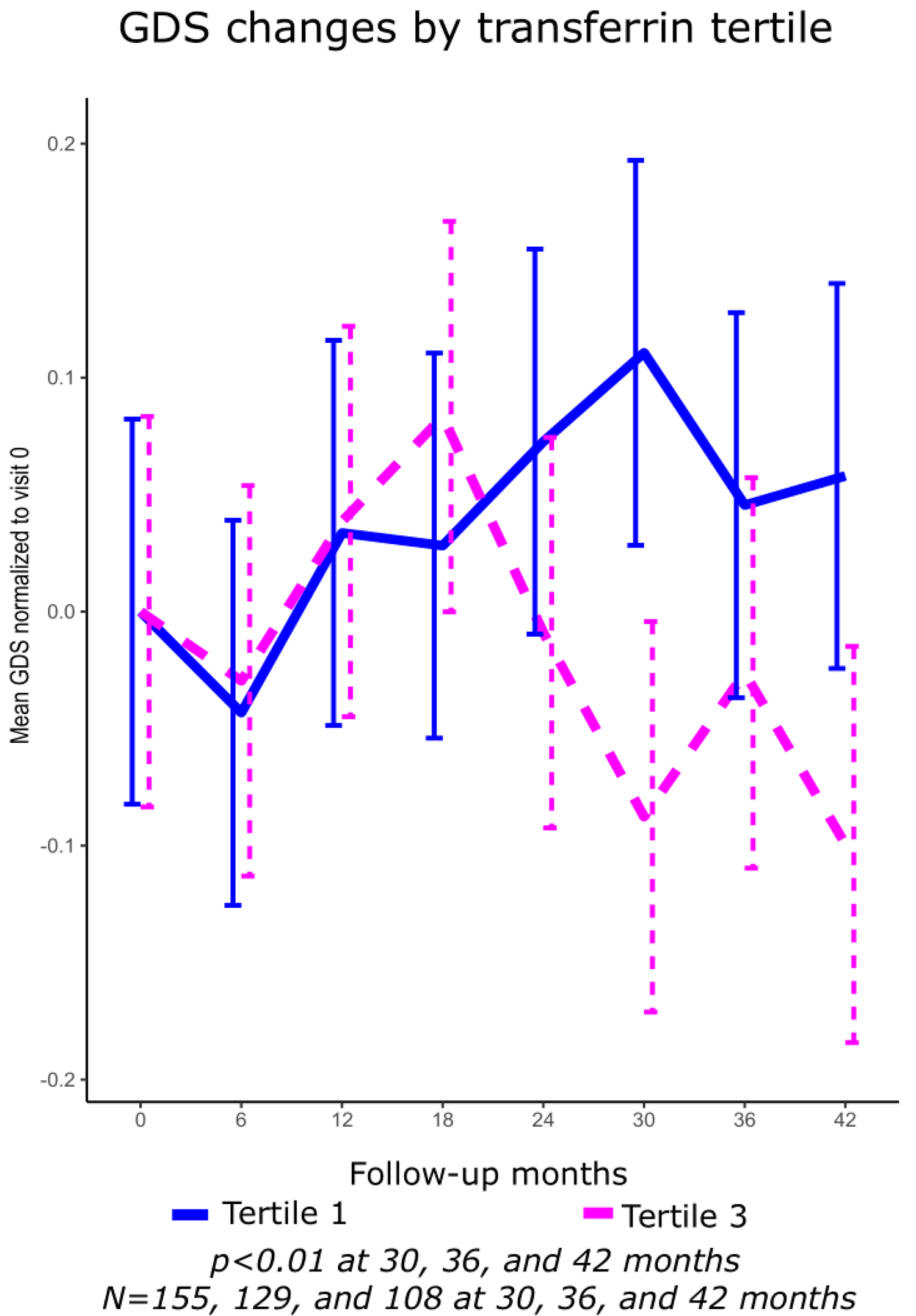
Changes in the Global Deficit Score (GDS) over time in study participants with CSF transferrin in tertiles 3 *vs*. 1 (*p*<0.01 for transferrin at 30, 36 and 42 months, adjusting for comorbidity, plasma HIV RNA, and zidovudine use).

(Associations remained unchanged even after repeating the analysis, excluding tertile 2, shown in **Supplemental Material, Figure S1**.)

As shown in **Supplemental material (Table S4**), associations with the GDS over time remained statistically significant at 36 and 42 months for H-ferritin in people with plasma viremia, but statistical significance was lost for transferrin associations in the virally suppressed subset. Both transferrin and H-ferritin longitudinal associations with the GDS also persisted after adjustment for CSF TNF-α and/or IL-6 levels. GDS changes over time in all three tertiles of H-ferritin and transferrin, normalized to baseline values, are presented in **Supplemental material** (**Figures S1** and **S2**).

### Age-stratified longitudinal observations

Since body iron stores and inflammation generally increase with age, and BBB integrity has been reported to decline with age [17], we determined whether the impact of CSF iron biomarkers on neurocognitive performance differed between younger and older people with HIV. The impact of CSF H-ferritin at baseline on GDS differences over time was pronounced in people with HIV under age 50 years, adjusting for comorbidity, a comorbidity interaction term, and plasma HIV RNA (uncorrected *p*-values 0.022, 0.004, 0.001, and Huynh-Feldt-corrected *p*-values 0.034, 0.013, 0.006 at 30, 36 and 42 months, respectively (**Supplemental material, Table S5** and **Figure S3**). The H-ferritin interaction with comorbidity showed borderline statistical significance at 42 months. Again, results were unchanged by additional adjustment for biomarkers of inflammation (either CSF TNF-α or IL-6). By contrast, CSF transferrin at baseline predicted differences in the GDS over time mainly in individuals aged 50 and older (uncorrected *p-*values 0.015, 0.039, 0.026; Huynh-Feldt-corrected *p*-values 0.038, 0.087, 0.078 at 30, 36, and 42 months, respectively; see **Figure S4**). Corrected *p-*values for a multiplicative transferrin-by-comorbidity interaction term remained statistically significant at 30 and 36 months.

Since neurocognitive change status at the last follow-up visit encompassed qualitatively different transitions in individual participants, we also compared CSF iron-biomarker levels between the following groups: 1) all participants whose neurocognitive performance improved from asymptomatic neurocognitive impairment (ANI) at baseline to *not impaired* at last follow-up visit *vs*. people who converted from ANI to either mild neurocognitive disorder (MND) or HIV-associated dementia (HAD); 2) participants who converted from *not impaired* to ANI at the last visit *vs*. individuals who improved from ANI at baseline to *not impaired* (**Supplemental material, Table S6**). (These comparisons captured clinically relevant transitions that might be missed in the neurocognitive *change* analyses described previously, which compared groups categorized regardless of impairment status as *improving, declining* or *stable* overall.) H-ferritin levels were approximately twice as high in individuals who converted to *not impaired* at 42 months than in people whose neurocognitive function declined further, though numbers in each group were small and the finding was only marginally statistically significant. CSF transferrin levels were also 2-3-fold higher in converters from ANI to *not impaired* than in people who made the opposite transition (*p*=0.08).

### Sensitivity analyses evaluating robustness of associations

We repeated longitudinal analyses at 42 months after 1) excluding biomarker outlier values in the entire cohort and in younger individuals (**Supplemental material, Table S7**), and 2) excluding tertile 2 data from ANOVA models (**Supplemental material, Table S8**). In addition, revised, practice-adjusted GDS values were provided by CHARTER investigators after the original analyses were performed, which altered baseline GDS values for 10% of study participants, so we performed our analyses using these updated variables. The observed associations were robust, as results of all of these analyses were essentially unchanged. Finally, a comparison of demographic and HIV disease characteristics between all CHARTER Study participants who were followed longitudinally for 30 months or more (evaluated in the present study) *vs*. participants who were lost to follow-up after the baseline assessment (not evaluable), is presented in **Supplemental material, Table S9**. Losses to follow-up included more individuals with undetectable plasma HIV RNA (*p*=0.039).

## DISCUSSION

Iron dysregulation has long been linked to neurodegenerative disorders, although it has been challenging to distinguish bystander effects from involvement in disease pathogenesis. Prospective studies evaluating the role of iron and iron-delivery proteins in HAND/NCI have not been performed previously. This also represents the first study, of which we are aware, to comprehensively evaluate multiple complementary indices of CNS iron status in CSF in a large number of adults with HIV, and to study levels of CSF heavy-chain (H)-ferritin in relation to longitudinal neurocognitive performance. Importantly, this study is one of the first to identify CSF biomarkers of neurocognitive *improvement* in people with HIV, a previously highlighted research priority, since NCI in adults with HIV on ART is often non-progressive [1, 18-19]. Prior studies were anecdotal or evaluated serum or CSF total ferritin only, and HIV-seropositive participants were either ART-naïve or had interrupted ART [17]. Serum ferritin is composed primarily of light-chain (L) subunits, which, more so than H-ferritin, are potently induced by inflammation [13]. In a small pre-ART-era study, Deisenhammer *et al* [20] reported elevated CSF ferritin in people with HIV who had diverse neurological conditions, including HAD. In contrast, this study assessed the independent contributions of H-ferritin, transferrin, and iron to NCI in over 400 adults with HIV on ART with detailed medical, neurocognitive, and neuropsychiatric follow-up for up to 42 months. H-ferritin contains all of the ferroxidase activity of the ferritin molecule, which is essential for its iron-scavenging and antioxidant properties [13]. In accord with prior published reports [21], CSF H-ferritin and transferrin were not correlated with contemporaneously measured CSF TNF-α or IL-6 in our study, nor did additional adjustment for CSF inflammatory cytokine levels alter the observed iron-biomarker associations with GDS differences over time. These observations indicate that, rather than reflecting inflammation, H-ferritin and transferrin perform important iron-homeostatic and antioxidant functions in the CNS. Significant and robust associations of higher CSF H-ferritin and transferrin at baseline with *improving* and particularly with better GDS-defined neurocognitive function over multiple follow-up visits are consistent with novel neuroprotective and/or neurotrophic roles for these antioxidant iron-transport proteins in the brain. Furthermore, the pattern of associations differed significantly for H-ferritin and transferrin, in that CSF H-ferritin predicted GDS differences over time in (and were strongest in) virally suppressed persons on ART, in younger adults with HIV, and in individuals with minimal comorbidity, whereas CSF transferrin associations with the GDS were mainly among older and/or viremic adults. Higher levels of both biomarkers were generally associated with better neurocognitive function at 30-42 months of follow-up.

Iron is critical for mitochondrial function, as well as HIV replication, but excess tissue iron and non-transferrin-bound (oxidatively active) iron (NTBI) also induce oxidative damage to cell membrane lipids and other structures. Moreover, inflammation increases the levels NTBI and NTBI transport across the BBB [22]. Neurons and astrocytes derive essential iron via receptor-mediated endocytosis of transferrin-bound iron and to a lesser extent, from NTBI present in brain interstitial fluid [22]. Myelinating oligodendrocytes lack transferrin receptors, however, and derive iron from H-ferritin via the Tim-1 receptor; H-ferritin released from microglia is trophic for these cells [12]. Recent data from our group raise the possibility that H-ferritin could also prevent oligodendrocyte toxicity due to Tim-1 binding to semaphorin 4A, levels of which are elevated in people with HIV [23]. Damage to white matter, which consists of myelinated nerve fibers, is an early and prominent finding in adults with HIV and correlates with NCI [24]. HIV infection may therefore promote functional iron deficiency due to HIV-mediated disruption of iron transport [15, 25], oxidative injury, or both, in the brain. Indeed, iron deficiency during early development has been associated with abnormally low brain iron content and with persistently impaired cognition later in childhood [16, 26]. Recently, Larsen et al. reported an increasing association between higher regional brain iron content, estimated using R2* relaxometry neuroimaging, and superior cognitive ability in several domains through late adolescence and young adulthood [16]. H-ferritin and transferrin, which maintain, transport, and deliver iron to metabolically active cells in a non-reactive state, are therefore positioned to play key roles in brain health, particularly during HIV infection. H-ferritin binds up to several-thousand-fold more iron atoms than other iron-binding proteins, including transferrin; via this iron-scavenging activity, it provides a significant buffer against iron-mediated oxidative stress and can also efficiently deliver iron when needed [13]. Altered levels of ferritin or ferritin subunits are reported to occur in several neurodegenerative disorders [27]. Higher CSF H-ferritin may offset oxidative injury caused by inflammation-related increases in NTBI and heme oxygenase-1 (HO-1) deficiency in the setting of HIV [28]. Intriguingly, H-ferritin mediates the antioxidant effect of HO-1 and histone deacetylase inhibitors and has emerging proteostatic, mitochondria-protective, immune-modulating, and signaling roles [29, 30]. Transferrin, particularly apo-transferrin, also confers neuroprotection by supporting myelination, reducing mitochondria-mediated cell death and upregulating cell-survival pathways, independent of its iron delivery functions [31, 32]. We therefore speculate that H-ferritin and transferrin, produced at least partly within the CNS and regulated by glia [12, 17], act to preserve iron bioavailability to myelinating oligodendrocytes and neurons, respectively, during HIV-or comorbidity-related inflammation, while limiting mitochondrial dysfunction and oxidative stress in the brain.

CSF is sequestered behind blood-brain and blood-CSF-barriers, and iron indices in CSF correlate poorly with serum levels, supporting the concept that they are actively regulated within the CNS compartment [17]. Higher CSF H-ferritin and transferrin could therefore indicate either decreased or increased bioavailable brain iron [15]. If brain iron and NTBI levels are increased due to inflammation and a compromised BBB, H-ferritin would be expected to protect against iron-mediated toxicity by acting as an iron sink [13, 29]. While higher CSF H-ferritin and transferrin levels could possibly reflect reduced transport of these proteins across the BBB, the evidence supporting H-ferritin and transferrin regulation within the CNS, and reported increases in BBB permeability in HIV infection, argue against this scenario [17, 33]. Most CSF transferrin is thought to be produced by the choroid plexus, which itself expresses H-ferritin and high levels of iron-transport proteins and has been shown to respond to systemic inflammation by altering its expression of iron-related genes [34, 35]. Additional studies are needed to determine the sources of CSF H-ferritin and transferrin in CSF and how CSF levels of these proteins relate to levels of these proteins in the brain and to brain iron in people with HIV.

Published data supporting beneficial effects of H-ferritin in the brain include reports that H-ferritin-mediated CXCR4 inhibition may reduce glutamatergic toxicity [36], and that mice engineered to overexpress H-ferritin in the brain show reduced glial activation and energy metabolism shifted towards anaerobic glycolysis, with increased lactate production [37]. Increased anaerobic glycolysis in the CNS has been previously associated with improving neurocognitive performance in people with HIV [38]. Either H-ferritin-deficient or H-ferritin-overexpressing mice may show evidence of iron deficiency in the brain, and the former also exhibit increased oxidative stress [29, 39, 40]. The beneficial effects of H-ferritin that we found in people with HIV may, however, be outweighed by iron-mediated oxidative stress in the setting of opioid drug abuse, which may increase endolysosomal iron (accompanied by neuronal H-ferritin upregulation) and dendritic spine loss [41]. Since the effect of transferrin was primarily in individuals with viremia, it is possible that this protein protects against oxidative stress by reducing NTBI entry into the brain during inflammation and preserving the iron supply to neurons and immature or differentiating oligodendrocytes, which depend upon transferrin-bound iron. Low levels of a transferrin homolog, melanotransferrin, in CSF have similarly been linked to increased risk of conversion from mild cognitive impairment to Alzheimer’s dementia [42]. Further studies are needed to clarify the interactions we observed between comorbidity and these iron-transport proteins: levels of systemic inflammation may have been higher in individuals with mild-to-moderate comorbidity in this sample and people with plasma viremia, and H-ferritin and transferrin may play distinct neuroprotective roles under scenarios of absent or minimal *vs*. active inflammation, respectively.

The impact of HIV and ART on iron delivery and iron metabolism in the human brain may be relevant to the persistence of HAND into the current ART era: brain iron is of emerging importance to cognitive development in children and young adults perinatally infected with HIV, who currently require lifelong ART [16]. Recent studies also suggest that postnatal brain acquisition of iron may differ significantly by sex, with males relying more on H-ferritin and females more on transferrin, for brain iron uptake [43]. To our knowledge, this is the largest study to date involving measurement of CSF iron indices in people with HIV; nevertheless, it was limited by a modest sample size and insufficient numbers of participants over age 60 to fully investigate age-related interactions of iron with *APOE*-*ε4* genotype on neurocognitive function. Although our analyses relied on single rather than serial CSF iron-biomarker measurements, normalization of GDS values at follow-up to baseline values within individuals, and the lengthy follow-up duration, mitigate concerns about sampling error or misclassification of neurocognitive impairment. The consistent direction of associations of CSF H-ferritin and transferrin with GDS-defined neurocognitive performance over 30, 36 and 42 months of follow-up, despite declining sample sizes at later visits, are all the more compelling given the single measurement at baseline. These findings further suggest the validity and significance of the neuroprotective effects we observed, which may take time to manifest (*e*.*g*., changes in myelination). Furthermore, associations were unchanged following a variety of sensitivity analyses, including re-analysis of the data using GDS values that were independently revised by CHARTER Study investigators in 10% of study participants, and following exclusion of outlier biomarker values, suggesting robust effects. Finally, while it would have been ideal to include serum biomarker or CSF:serum albumin ratio data as covariates in our analyses, these data were not available, and adjustment of models for age and plasma HIV RNA - two major factors that influence BBB integrity – did not alter the observed associations [17]. Taken together, these results support the validity of the observed associations.

In conclusion, results from this prospective study suggest that higher CSF H-ferritin and transferrin reflect improving brain iron homeostasis and predict significantly better neurocognitive performance over time in adults with HIV. The benefits of higher H-ferritin extend to younger and importantly, to virally suppressed individuals, while higher transferrin levels are primarily protective in older people with HIV and in viremic individuals, who likely have increased levels of inflammation. Future large-scale studies that include larger numbers of older as well as HIV-seronegative individuals may clarify the relative contributions of HIV and aging to iron dysregulation in the brain. The novel findings from this study provide hope that novel iron-modulating interventions such as H-ferritin might be developed to prevent NCI in adults with HIV, or to halt its progression.

## METHODS

### Study design and participants

The U.S. CNS HIV Anti-Retroviral Therapy Effects Research (CHARTER) Study is a prospective, multi-center observational study of neuro-HIV outcomes [44]. The CHARTER Study was approved by the Human Subjects Protection Committees of each participating institution, and all participants provided written informed consent. Participants were selected from the CHARTER/National NeuroAIDS Tissue Consortium (NNTC) biospecimen repository based on availability of longitudinal assessment data and stored CSF, collected by lumbar puncture, from the baseline (entry) visit. CHARTER Study protocols adhere to the ethical principles of the Helsinki Declaration.

### Assessment of HAND/NCI

CHARTER Study participants without severe comorbidity (such as a learning disability, epilepsy, or traumatic head injury with prolonged loss of consciousness) that could confound a diagnosis of HAND, as previously described [45], underwent comprehensive neurocognitive testing at baseline and follow-up visits 6 months apart. The battery included 15 tests, which assess 7 ability domains, allowing for calculation of a GDS. The GDS incorporates up-to-date norms for age and other demographics, and corrections for learning/practice effects [45]. Neurocognitive outcomes evaluated in this study included the GDS as either a continuous variable, or a dichotomous variable, using an established cut-off of GDS ≥0.50 to define NCI [46]. Frascati criteria for HAND were used to classify individuals as having *no impairment*, ANI, MND, or HAD [46]. Neurocognitive change status was categorized as *improving, stable*, or *declining* at each follow-up visit compared to baseline and was determined from a z-score generated for all 15 neuropsychological tests, based on published normative data [45]. Neurocognitive change status at the last follow-up was defined as 1) *declining*: if a participant had at least one “declined” status but no “improved” status at any visit(s); 2) *improving*: if a participant had at least one “improved” status and no “declined” status; 3) *stable*: if a participant had unchanged neurocognitive status at all visits [44]. Psychiatric and substance-use diagnoses were determined as reported previously [45].

### Quantification of CSF iron biomarkers and inflammation

CSF iron-biomarker levels were determined using commercially available assay kits according to the manufacturer’s protocol [17]. Concentrations of inflammatory cytokines and chemokines were measured in CSF by commercial, high-sensitivity multiplex assay [interleukin-6 (IL-6), tumor-necrosis-α (TNF-α)], according to the supplier’s protocol (Luminex FlexMap 3D platform, Millipore, Billerica, MA). All biomarkers were measured within a 4-5-hour time window to avoid significant diurnal differences. Iron and iron-carrier proteins have been measured routinely for many years, and their stability and solubility in stored human biospecimens is well established [40, 47-50]. Laboratory personnel who performed the assays were unaware of participant clinical outcomes.

### Statistical Methods

The GDS as a continuous variable was compared across baseline demographic and HIV disease factors using either analysis of variance (factors or variables with two or more categories), linear regression (normally distributed continuous variables), or the Wilcoxon Rank Sum test (non-normally distributed variables). Pearson’s *r*-values were used to assess iron-biomarker and inflammation-biomarker correlations. The multivariable analyses performed and participant subsets included in each analysis are summarized in **Figure 4**. Associations between neurocognitive change status and CSF iron biomarkers at baseline were evaluated by logistic regression analysis, adjusting for age (<50 *vs*. ≥50 years), comorbidity severity (none/minimal *vs*. mild-to-moderate), plasma HIV RNA (≤50 *vs*. >50 copies/mL), and *APOE-*ε*4* allele status (carrier *vs*. non-carrier; genotype distributions summarized in **Supplemental material, Table S1**). While the GDS incorporates some age adjustment, age is a strong risk factor for NCI and was included in models to reduce residual confounding. Since use of the antiretroviral drug zidovudine was previously associated with CSF transferrin in this sample, zidovudine use was also included as a covariate in analyses of transferrin [2]. Odds ratios (ORs) for *improving vs. stable, declining vs. stable*, and *improving vs. declining* neurocognitive change status at the last follow-up assessment, and their 95% CIs, were estimated, adjusting for age, comorbidity, *APOE-ε4* carrier status, and plasma HIV RNA, as well as zidovudine in transferrin analyses.

**Figure 4.**
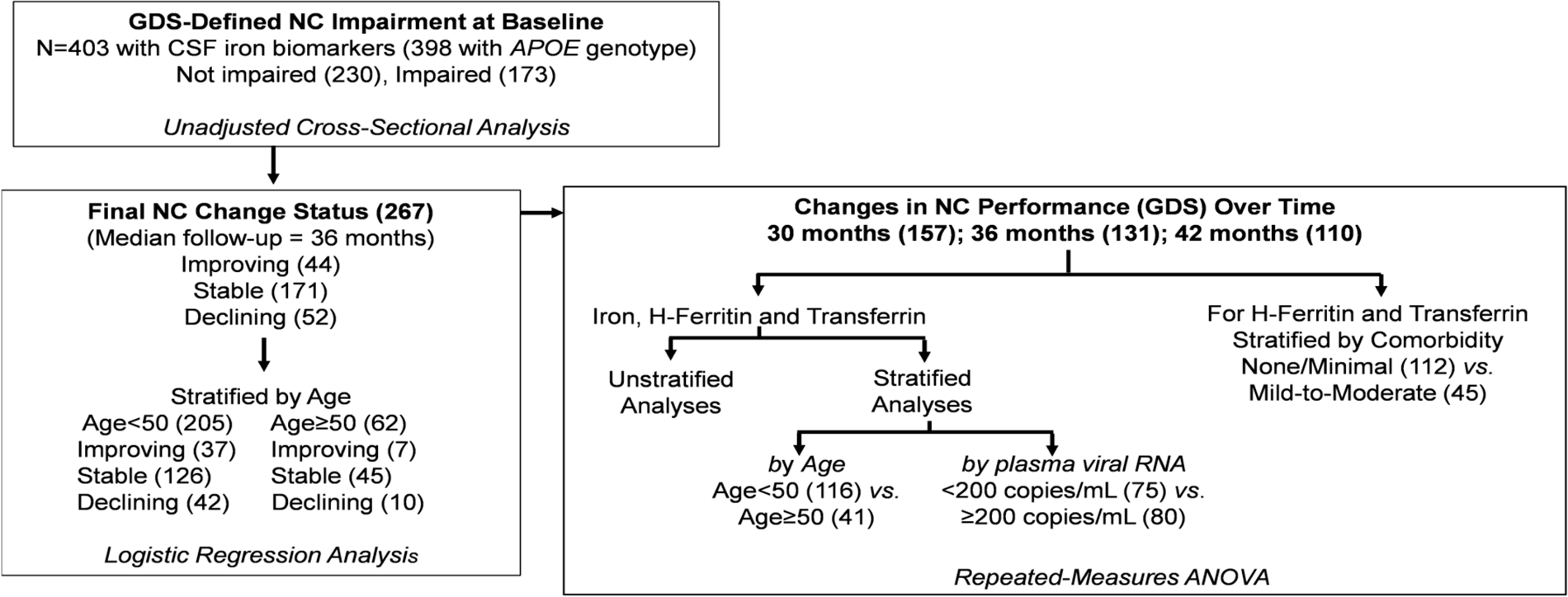
Study schema showing numbers of participants (in parentheses) who were included in individual analyses. *Abbreviations: CSF*, Cerebrospinal fluid; *GDS*, Global Deficit Score; *NC*, Neurocognitive; *ANI*, Asymptomatic Neucocognitive Impairment; *MND*, Mild Neurocognitive Disorder; *HAD*, HIV-Associated Dementia; *ANOVA*, Analysis of variance.

We performed repeated-measures analysis of variance (ANOVA) regression to evaluate the influence of all three iron-related biomarkers (indicating CSF iron status at baseline) on differences in GDS-defined neurocognitive function over 30, 36, and 42 months of follow-up, normalized to baseline values. In these longitudinal analyses, CSF iron biomarkers were categorized into tertiles to facilitate interpretation. The *p*-values at specific follow-up visits reflect differences in the GDS between the highest and lowest biomarker tertiles, summarized over all visits up to and including that visit; however, all three biomarker tertiles were included in ANOVA regression analyses. Effect sizes for associations were estimated using the *eta*-squared (η^2^) statistic, interpreted as the proportion of total variance in the dependent variable (GDS) that is accounted for by membership in a biomarker tertile. All models were adjusted for comorbidity and plasma HIV RNA. Models additionally adjusted for *APOE-*ε*4* carrier status, and models stratified by age (<50 *vs*. ≥50 years), comorbidity severity, and plasma HIV RNA (viral suppression, <200 copies/mL *vs*. ≥200/mL), were also evaluated. This age cut-off was chosen for stratification, due to low numbers of study participants 60 years of age and older (the age group in which *APOE-*ε*4* associations with NCI were previously reported) [9]; the higher plasma HIV RNA cut-off was used in this case in order to maximize power in the strata being analyzed.

As iron homeostasis in HAND is a new area of investigation, and we previously associated CSF H-ferritin and transferrin with comorbidity and zidovudine use, we also stratified H-ferritin and transferrin models by comorbidity and zidovudine use, respectively, and tested for multiplicative interactions between these variables [17]. Adjustment for self-reported race/ethnicity did not significantly alter results, and the GDS includes corrections for demographic norms, so race/ethnicity was not included in the final models. Since Mauchly’s test of the sphericity assumption for repeated-measures ANOVA was statistically significant, we also present ANOVA results with *p-*values that incorporate the Huynh-Feldt *epsilon* correction [51]. This correction minimizes the Type I error rate but is not overly stringent or biased for smaller samples.

Importantly, to evaluate the impact of inflammation, CSF TNF-α level and/or IL-6 level at baseline was included as a covariate in separate multivariable logistic or ANOVA regression models. Results including these biomarkers were essentially unchanged. ANOVA regression models were adjusted for CSF TNF-α (measured at baseline) at the 30-, 36-, and 42-month visits to ensure that longitudinal associations with iron biomarkers were not confounded by inflammation. Robustness of associations was also evaluated by excluding iron-biomarker outlier values 2 or more standard deviations above or below the mean. CHARTER Study investigators also re-calculated baseline GDS values to improve upon adjustments for practice effects that might have occurred (learning that may occur when an individual is assessed more than once using the same instrument) and revised 10% of GDS values; we therefore repeated all analyses using the new data. To evaluate potential selection bias in the data, HIV disease characteristics and CSF iron-biomarker levels at baseline were compared between study participants who were followed for the full 42 months and individuals who were lost to follow-up after 30 months.

Finally, we compared CSF iron biomarkers at baseline between groups of study participants whose neurocognitive status converted as follows: 1) *not impaired* at baseline to ANI at last follow-up *vs*. ANI at baseline to *not impaired* at last follow-up, and 2) ANI at baseline to *not impaired* at last follow-up *vs*. ANI at baseline to either MND or HAD at the last assessment. These analyses were adjusted for comorbidity severity and zidovudine use.

Statistical significance for all analyses was set at a two-tailed *p*-value<0.05. Statistical analyses were conducted using STATA Statistical Software, Release 13 (College Station, Texas: StataCorp LP).

## Data Availability

The dataset analyzed for this manuscript is available upon request.

## Acknowledgments

We are grateful to all of the individuals who participated in this study. We also acknowledge the efforts of other CHARTER Study Group members who assisted in recruitment at participating study sites:

## Other CHARTER Study Group members

In addition to the authors, the CHARTER study is affiliated with the following six participating study sites: Johns Hopkins University; the Icahn School of Medicine at Mount Sinai; University of California, San Diego; University of Texas, Galveston; University of Washington, Seattle; and Washington University, St. Louis. CHARTER is headquartered at the University of California, San Diego and includes: Director: Igor Grant, M.D.; Co-Directors: Scott L. Letendre, M.D., Ronald J. Ellis, M.D., Ph.D., Thomas D. Marcotte, Ph.D.; Center Manager: Donald Franklin, Jr.; Neuromedical Component: Ronald J. Ellis, M.D., Ph.D. (P.I.), J. Allen McCutchan, M.D.; Laboratory and Virology Component: Scott Letendre, M.D. (Co-P.I.), Davey M. Smith, M.D. (Co-P.I.).; Neurobehavioral Component: Robert K. Heaton, Ph.D. (P.I.), J. Hampton Atkinson, M.D., Matthew Dawson; Imaging Component: Christine Fennema-Notestine, Ph.D. (P.I.), Michael J Taylor, Ph.D., Rebecca Theilmann, Ph.D.; Data Management Component: Anthony C. Gamst, Ph.D. (P.I.), Clint Cushman; Statistics Component: Ian Abramson, Ph.D. (P.I.), Florin Vaida, Ph.D.; Johns Hopkins University Site: Ned Sacktor (P.I.), Vincent Rogalski; Icahn School of Medicine at Mount Sinai Site: Susan Morgello, M.D. (Co-P.I.) and David Simpson, M.D. (Co-P.I.), Letty Mintz, N.P.; University of California, San Diego Site: J. Allen McCutchan, M.D. (P.I.); University of Washington, Seattle Site: Ann Collier, M.D. (Co-P.I.) and Christina Marra, M.D. (Co-P.I.), Sher Storey, PA-C.; University of Texas, Galveston Site: Benjamin Gelman, M.D., Ph.D. (P.I.), Eleanor Head, R.N., B.S.N.; and Washington University, St. Louis Site: David Clifford, M.D. (P.I.), Muhammad Al-Lozi, M.D., Mengesha Teshome, M.D.

## Supporting information

**S1 Fig.**
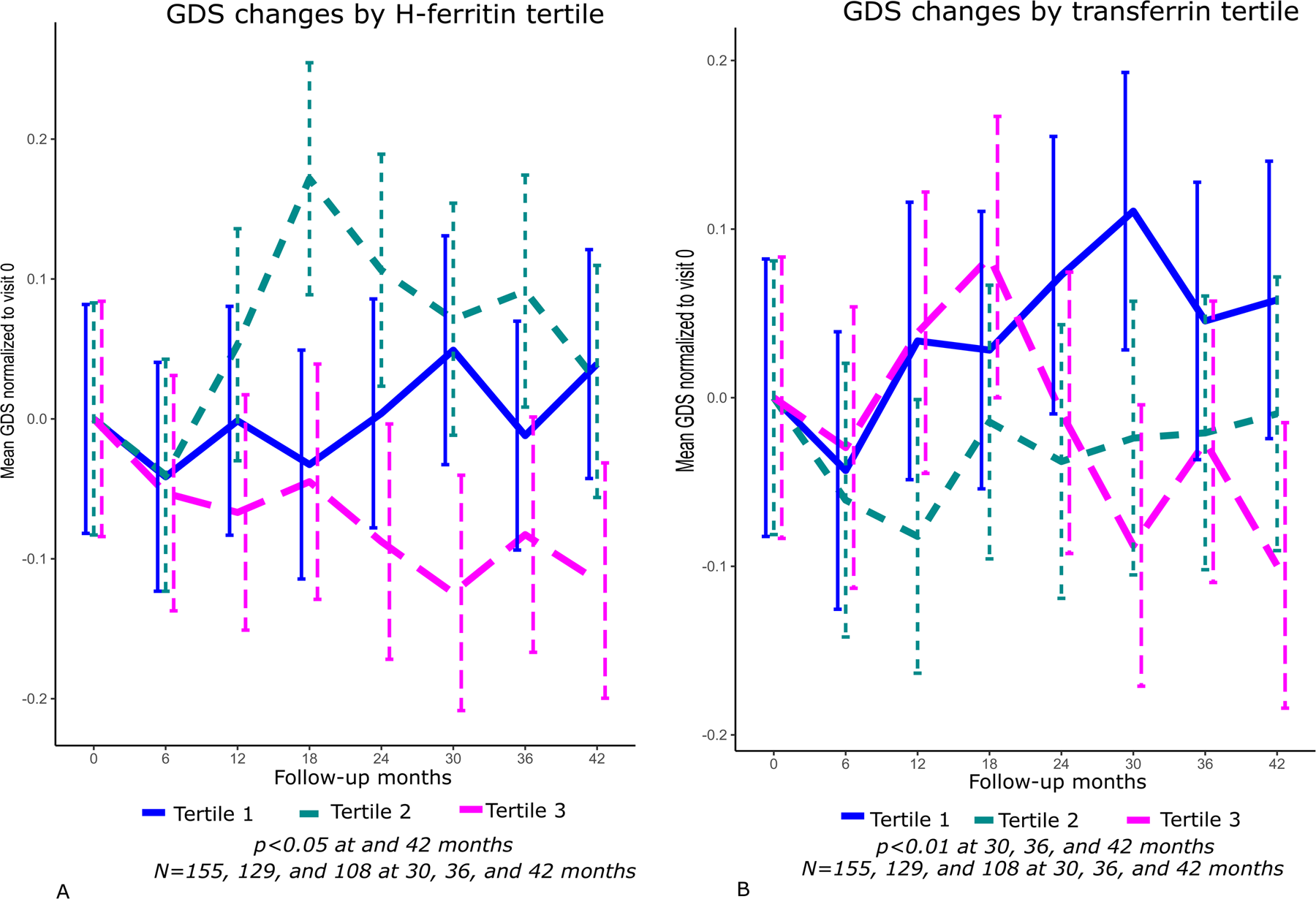
Changes in neurocognitive performance in CHARTER study participants, as measured by the Global Deficit Score (GDS) over time, in all three tertiles at baseline of CSF H-ferritin and transferrin (**panels *A*** and ***B***, respectively; both *p*<0.05 for tertiles 3 (pink) *vs*. 1 (blue) of H-ferritin over at 30 and 42 months (adjusting for comorbidity and plasma HIV RNA), and all *p*<0.01 for tertiles 3 *vs*. 1 of CSF transferrin at 30, 36, and 42 months, adjusting for comorbidity, zidovudine use and plasma HIV RNA). (PDF)

**S2 Fig.**
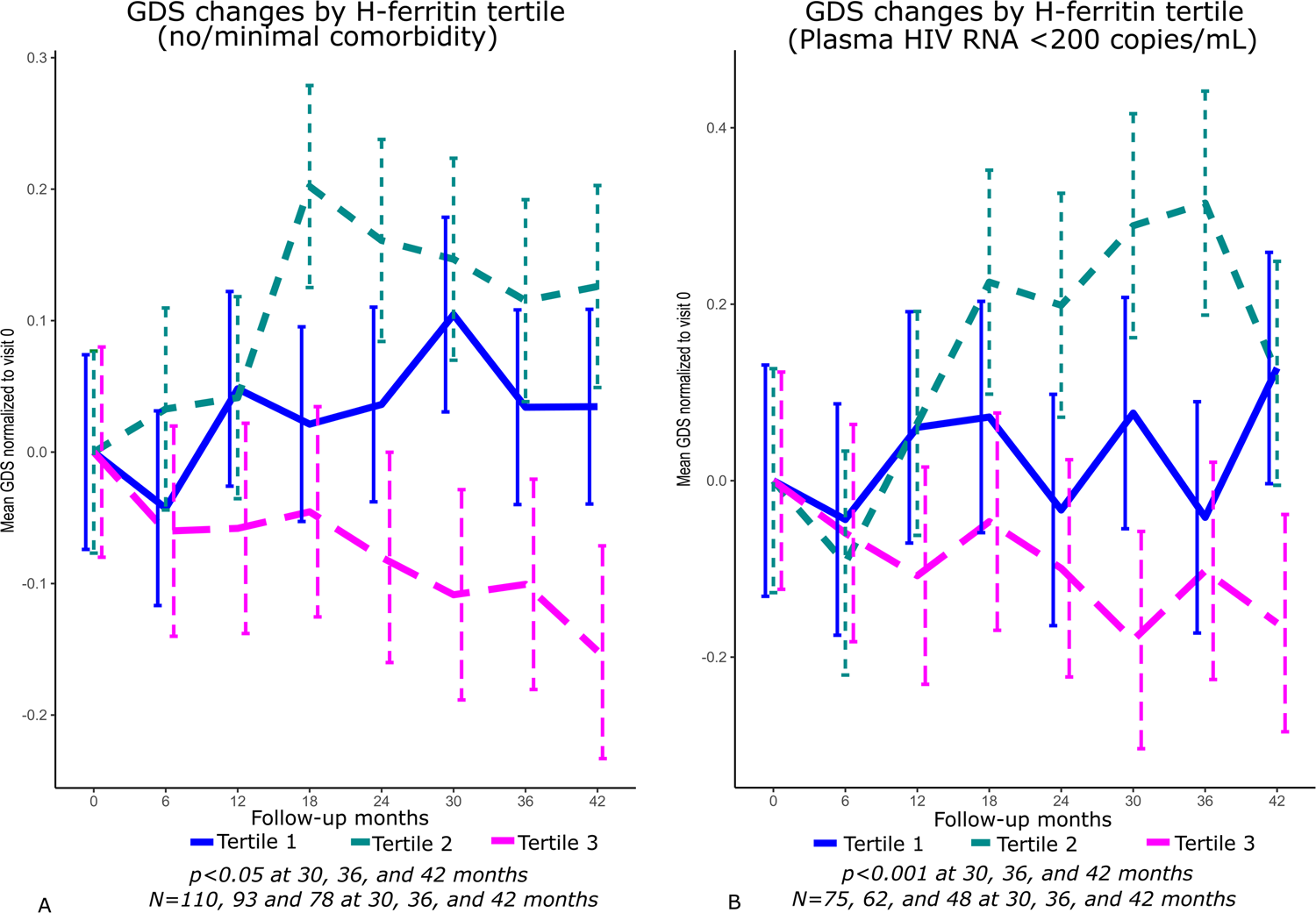
Changes in GDS over time, in all three tertiles at CSF H-ferritin among individuals with no/minimal comorbidity (**panel *A***, *p*<0.05 at 30, 36, and 42 months) adjusting for plasma HIV RNA and in individuals who were virally suppressed (**panel *B***, *p<0*.*001* at 30, 36, and 42 months of follow-up), adjusting for comorbidity. (PDF)

**S3 Fig.**
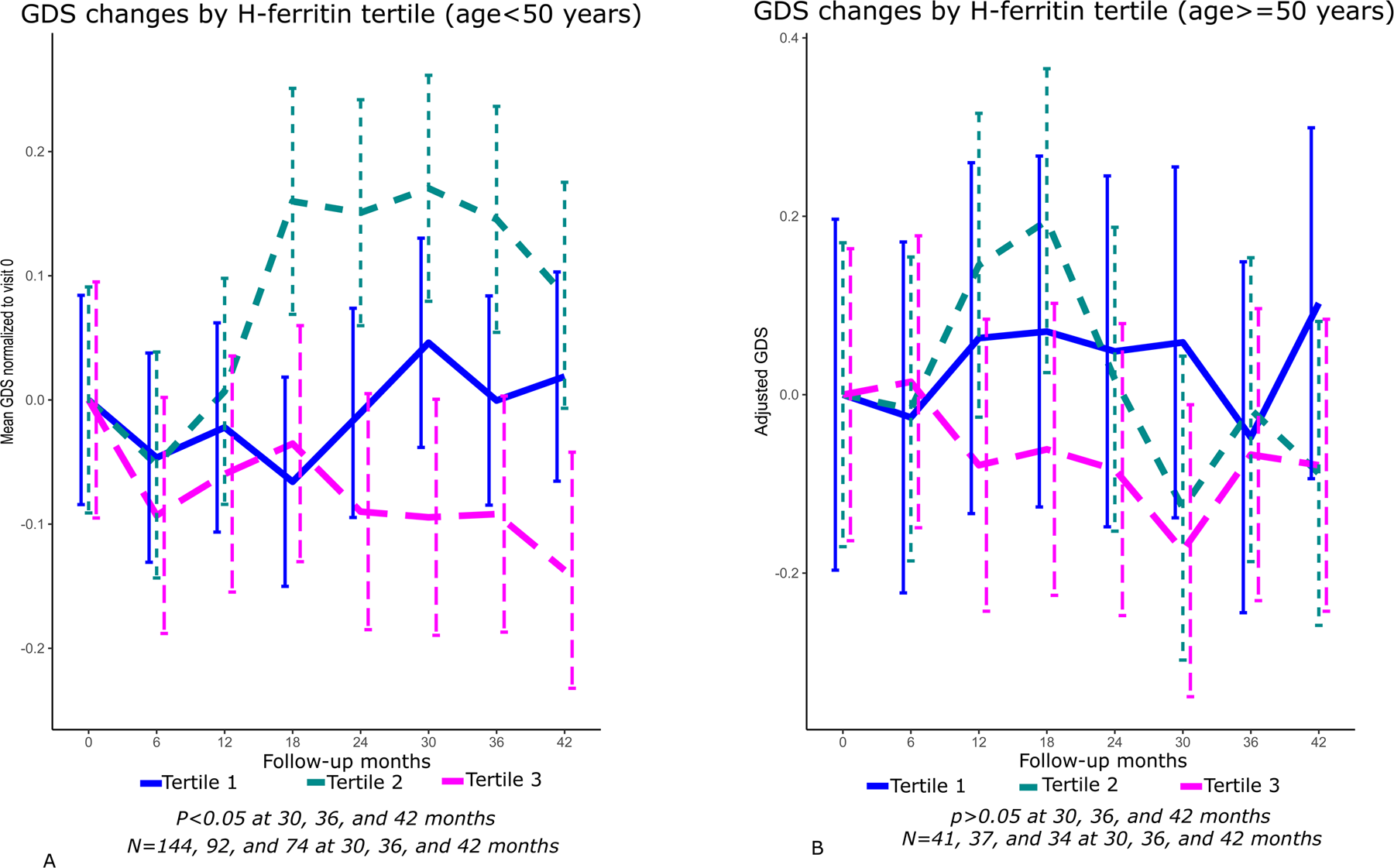
Results for CSF H-ferritin in individuals Panel A) aged <50 years are shown here (*p*<0.05 at 30 months and <0.01 for all subsequent visits for GDS differences in tertile 3 *vs*. 1, adjusting for comorbidity severity and plasma HIV RNA). Panel B) shows baseline CSF H-ferritin levels with GDS differences over 30, 36 and 42 months of follow-up in older individuals (p>0.05 at 30, 36, and 42 months). (PDF)

**S4 Fig.**
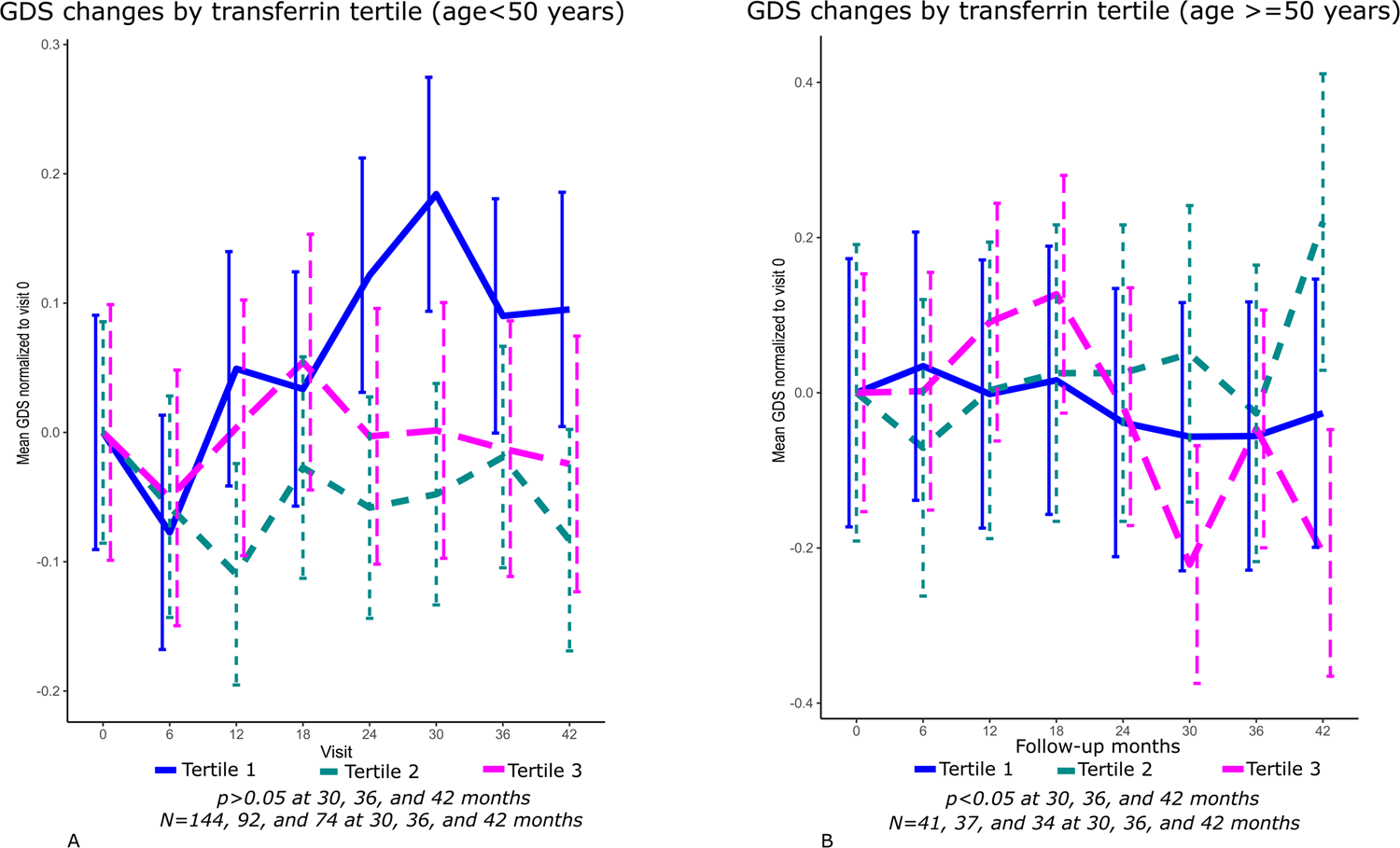
Changes in the GDS over time in all three tertiles of CSF transferrin at baseline among people with HIV, aged <50 years (**panel *A***, *p*=n.s. at all three time points) and aged ≥ 50 years (**panel *B***, *p*<0.05 at 30 and 42 months of follow-up) adjusting for comorbidity, plasma HIV RNA, and zidovudine use. (PDF)

**S1 Table.**
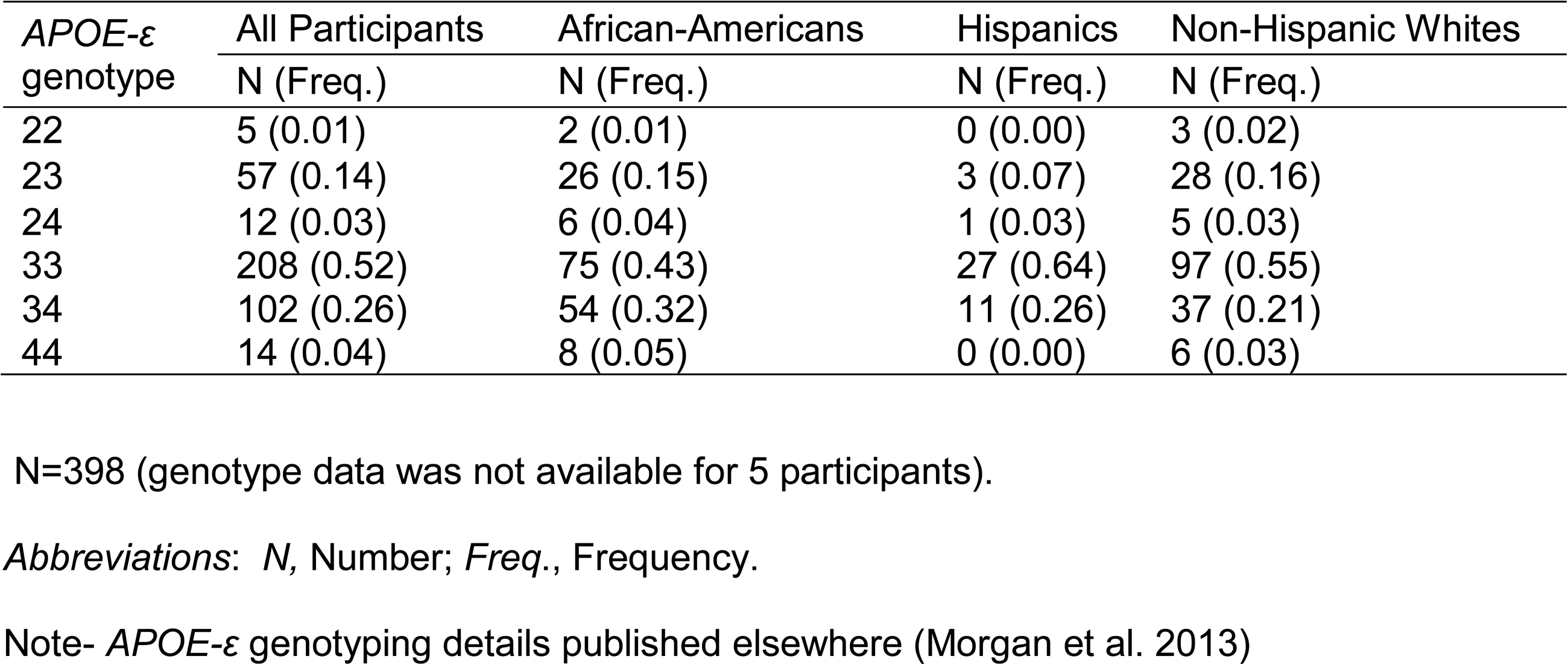
Overall *APOE* genotype frequency distribution in CHARTER Study participants by self-reported race and ethnicity.

| *APOE*-ε genotype | All Participants<br>N (Freq.) | African-Americans<br>N (Freq.) | Hispanics<br>N (Freq.) | Non-Hispanic Whites<br>N (Freq.) |
| --- | --- | --- | --- | --- |
| 22 | 5 (0.01) | 2 (0.01) | 0 (0.00) | 3 (0.02) |
| 23 | 57 (0.14) | 26 (0.15) | 3 (0.07) | 28 (0.16) |
| 24 | 12 (0.03) | 6 (0.04) | 1 (0.03) | 5 (0.03) |
| 33 | 208 (0.52) | 75 (0.43) | 27 (0.64) | 97 (0.55) |
| 34 | 102 (0.26) | 54 (0.32) | 11 (0.26) | 37 (0.21) |
| 44 | 14 (0.04) | 8 (0.05) | 0 (0.00) | 6 (0.03) |

N=398 (genotype data was not available for 5 participants).
*Abbreviations:* *N*, Number; *Freq.*, Frequency.
Note- *APOE*- $\epsilon$ genotyping details published elsewhere (Morgan et al. 2013)

**S2 Table.** Association of CSF H-ferritin at baseline with neurocognitive *change* status, compared to baseline, at the last follow-up visit.

| Variables in Model | OR (95% CI) *Improving* vs. *Stable* NC Performance | *p* | OR (95% CI) *Declining* vs. *Stable* NC Performance | *p* |
| --- | --- | --- | --- | --- |
| H-Ferritin | 1.12 (1.01 - 1.25) | **0.04** | 1.02 (0.91 - 1.15) | 0.72 |
| TNF-α | 0.86 (0.50 - 1.47) | 0.57 | 0.94 (0.71 - 1.26) | 0.69 |
| Comorbidity | 1.43 (0.70 - 2.89) | 0.33 | 1.69 (0.88 - 3.24) | 0.12 |
| Age | 0.97 (0.93 - 1.01) | 0.19 | 0.96 (0.92 - 1.00) | **0.05** |

*N*=44, 171 and 52 for *improving*, *stable* and *declining* neurocognitive (NC) performance respectively.
*p*-values adjusting for a measured CSF biomarker of inflammation (TNF-α levels), comorbidity, and age at baseline (entry onto the study).
*Note:* The *stable* group here includes both neurocognitively impaired and unimpaired persons.
*p*-values<0.05 (**bolded**) are considered statistically significant.
Units of measurement- H-ferritin- μg/mL
*Abbreviations:* OR, odds ratio; TNF-α , tumor necrosis factor-*alpha*; NC, neurocognitive; CI, confidence interval; *p*, *p*-value.

**S3 Table.**
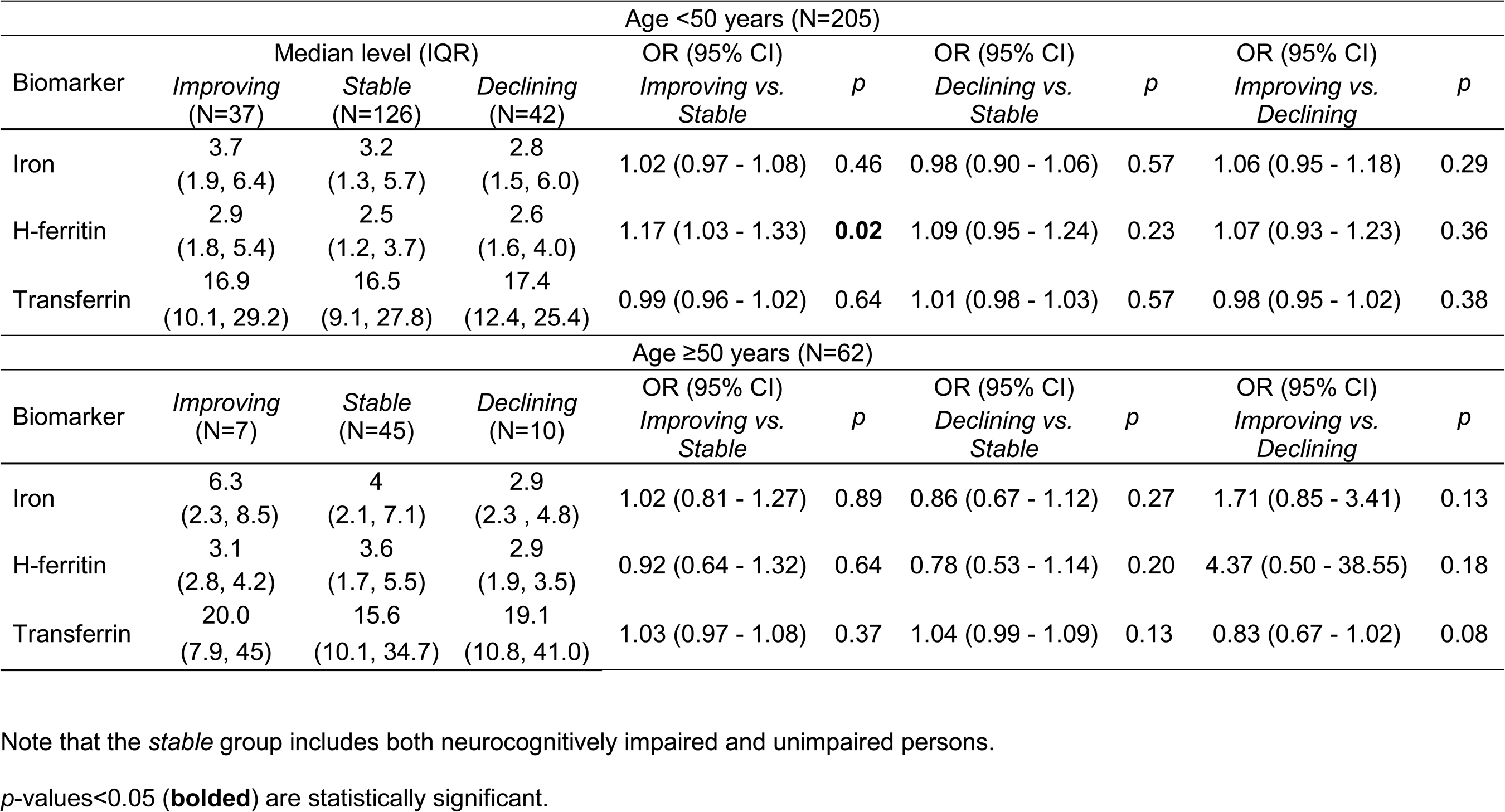
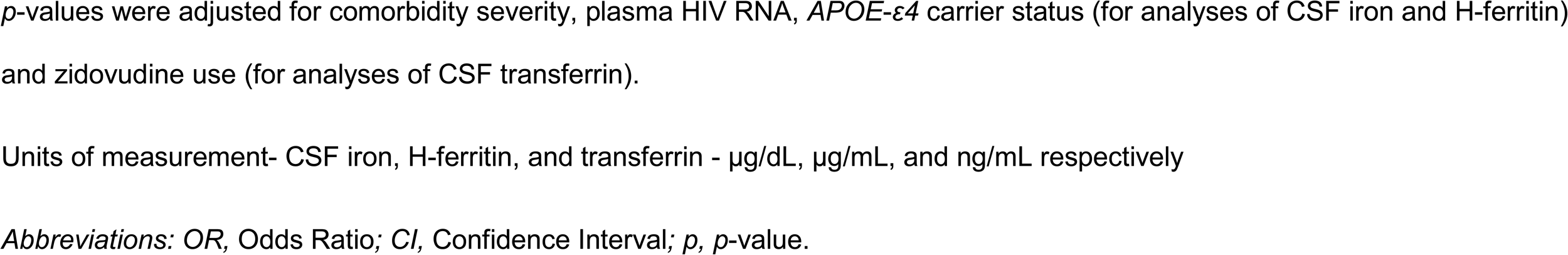
Distributions of CSF iron, H-ferritin, and transferrin levels at baseline among younger (<50 years) and older (≥50 years) neurocognitively *improving, stable* and *declining* people with HIV at their last follow-up visit.

| Age <50 years (N=205) |  |  |  |  |  |  |  |  |  |
| --- | --- | --- | --- | --- | --- | --- | --- | --- | --- |
| Biomarker | Median level (IQR) |  |  | OR (95% CI) |  | OR (95% CI) |  | OR (95% CI) |  |
|  | *Improving*<br>(N=37) | *Stable*<br>(N=126) | *Declining*<br>(N=42) | *Improving vs.*<br>*Stable* | *p* | *Declining vs.*<br>*Stable* | *p* | *Improving vs.*<br>*Declining* | *p* |
| Iron | 3.7<br>(1.9, 6.4) | 3.2<br>(1.3, 5.7) | 2.8<br>(1.5, 6.0) | 1.02 (0.97 - 1.08) | 0.46 | 0.98 (0.90 - 1.06) | 0.57 | 1.06 (0.95 - 1.18) | 0.29 |
| H-ferritin | 2.9<br>(1.8, 5.4) | 2.5<br>(1.2, 3.7) | 2.6<br>(1.6, 4.0) | 1.17 (1.03 - 1.33) | **0.02** | 1.09 (0.95 - 1.24) | 0.23 | 1.07 (0.93 - 1.23) | 0.36 |
| Transferrin | 16.9<br>(10.1, 29.2) | 16.5<br>(9.1, 27.8) | 17.4<br>(12.4, 25.4) | 0.99 (0.96 - 1.02) | 0.64 | 1.01 (0.98 - 1.03) | 0.57 | 0.98 (0.95 - 1.02) | 0.38 |
| Age ≥50 years (N=62) |  |  |  |  |  |  |  |  |  |
| Biomarker | Median level (IQR) |  |  | OR (95% CI) |  | OR (95% CI) |  | OR (95% CI) |  |
|  | *Improving*<br>(N=7) | *Stable*<br>(N=45) | *Declining*<br>(N=10) | *Improving vs.*<br>*Stable* | *p* | *Declining vs.*<br>*Stable* | *p* | *Improving vs.*<br>*Declining* | *p* |
| Iron | 6.3<br>(2.3, 8.5) | 4<br>(2.1, 7.1) | 2.9<br>(2.3, 4.8) | 1.02 (0.81 - 1.27) | 0.89 | 0.86 (0.67 - 1.12) | 0.27 | 1.71 (0.85 - 3.41) | 0.13 |
| H-ferritin | 3.1<br>(2.8, 4.2) | 3.6<br>(1.7, 5.5) | 2.9<br>(1.9, 3.5) | 0.92 (0.64 - 1.32) | 0.64 | 0.78 (0.53 - 1.14) | 0.20 | 4.37 (0.50 - 38.55) | 0.18 |
| Transferrin | 20.0<br>(7.9, 45) | 15.6<br>(10.1, 34.7) | 19.1<br>(10.8, 41.0) | 1.03 (0.97 - 1.08) | 0.37 | 1.04 (0.99 - 1.09) | 0.13 | 0.83 (0.67 - 1.02) | 0.08 |

Note that the *stable* group includes both neurocognitively impaired and unimpaired persons.
*p*-values<0.05 (**bolded**) are statistically significant.
*p*-values were adjusted for comorbidity severity, plasma HIV RNA, *APOE-ε4* carrier status (for analyses of CSF iron and H-ferritin) and zidovudine use (for analyses of CSF transferrin).
Units of measurement- CSF iron, H-ferritin, and transferrin - µg/dL, µg/mL, and ng/mL respectively
*Abbreviations: OR*, Odds Ratio; *CI*, Confidence Interval; *p*, *p*-value.

**S4-A Table.**
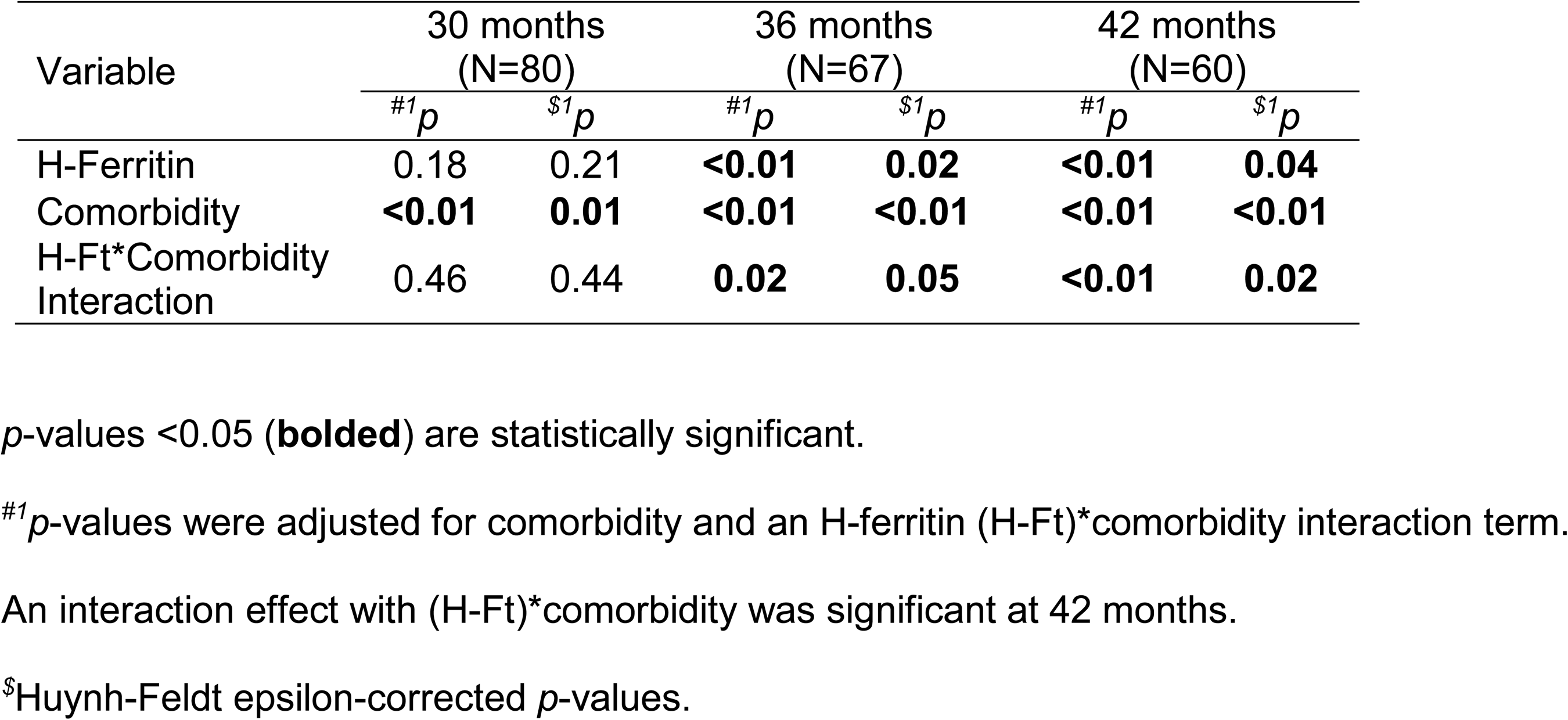
Associations of baseline CSF H-ferritin levels with GDS differences over 30, 36 and 42 months of follow-up in viremic adults with HIV (plasma HIV RNA ≥200 copies/mL).

| Variable | 30 months<br>(N=80) |  | 36 months<br>(N=67) |  | 42 months<br>(N=60) |  |
| --- | --- | --- | --- | --- | --- | --- |
| | <sup>#1</sup>*p* | <sup>\$1</sup>*p* | <sup>#1</sup>*p* | <sup>\$1</sup>*p* | <sup>#1</sup>*p* | <sup>\$1</sup>*p* |
| H-Ferritin | 0.18 | 0.21 | **<0.01** | **0.02** | **<0.01** | **0.04** |
| Comorbidity | **<0.01** | **0.01** | **<0.01** | **<0.01** | **<0.01** | **<0.01** |
| H-Ft*Comorbidity Interaction | 0.46 | 0.44 | **0.02** | **0.05** | **<0.01** | **0.02** |

*p*-values <0.05 (**bolded**) are statistically significant.
<sup>#1</sup>*p*-values were adjusted for comorbidity and an H-ferritin (H-Ft)\*comorbidity interaction term.
An interaction effect with (H-Ft)\*comorbidity was significant at 42 months.
<sup>\$</sup>Huynh-Feldt epsilon-corrected *p*-values.

**S4-B Table.** No significant associations of CSF transferrin with GDS differences in aviremic adults with HIV (plasma HIV RNA <200 copies/mL).

| Variable | 30 months (N=75) |  | 36 months (N=62) |  | 42 months (N=48) |  |
| --- | --- | --- | --- | --- | --- | --- |
| | <sup>#2</sup>*p* | <sup>\$2</sup>*p* | <sup>#2</sup>*p* | <sup>\$2</sup>*p* | <sup>#2</sup>*p* | <sup>\$2</sup>*p* |
| Transferrin | 0.49 | 0.47 | 0.68 | 0.60 | 0.48 | 0.45 |
| Comorbidity | 0.77 | 0.71 | 0.88 | 0.76 | 0.40 | 0.38 |
| TF*Comorbidity Interaction | 0.97 | 0.94 | 0.60 | 0.53 | 0.66 | 0.58 |
| Zidovudine use | **0.01** | --- | **0.02** | --- | 0.26 | --- |

*p*-values <0.05 (**bolded**) are statistically significant.
<sup>#2</sup>*p*-values were adjusted for comorbidity, zidovudine use and a transferrin (TF)\*comorbidity interaction term.
<sup>\$</sup>Huynh-Feldt epsilon-corrected *p*-values.

**S5-A Table.** Association of CSF H-ferritin with GDS differences over follow-up visits in younger adults (aged <50 years).

| Variable | 30 months (N=114) |  | 36 months (N=92) |  | 42 months (N=74) |  |
| --- | --- | --- | --- | --- | --- | --- |
| | <sup>#1</sup>*p* | <sup>1\$</sup>*p* | <sup>#1</sup>*p* | <sup>\$1</sup>*p* | <sup>#1</sup>*p* | <sup>\$1</sup>*p* |
| H-Ferritin | **0.02** | **0.03** | **<0.01** | **0.01** | **<0.01** | **<0.01** |
| Comorbidity | 0.92 | 0.88 | 0.87 | 0.81 | 0.91 | 0.84 |
| H-Ft*Comorbidity Interaction | 0.67 | 0.64 | 0.09 | 0.12 | **0.04** | 0.06 |
| Plasma HIV RNA | 0.24 | --- | 0.34 | --- | 0.64 | --- |

An interaction effect with comorbidity was borderline significant at 42 months.
*p*-values<0.05 (**bolded**) are statistically significant.
<sup>#1</sup>*p*-values were adjusted for comorbidity severity, plasma HIV RNA and an H-ferritin (H-Ft)\*comorbidity interaction term. Additional adjustment for *APOE-ε4* carrier status, CSF TNF-α, or IL-6 levels at baseline did not alter significant *p*-values for H-ferritin in the model (all *p*-values<0.05).
<sup>\$</sup>Huynh-Feldt epsilon-corrected *p*-values.

**S5-B Table.**
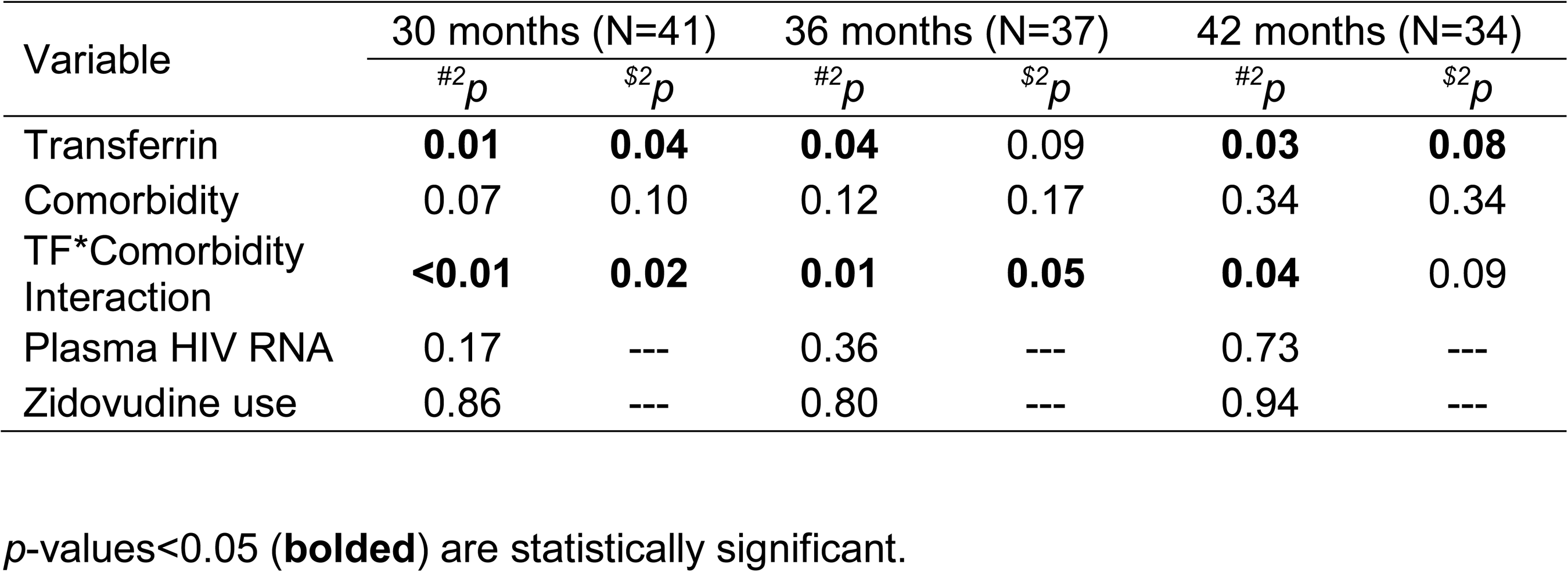
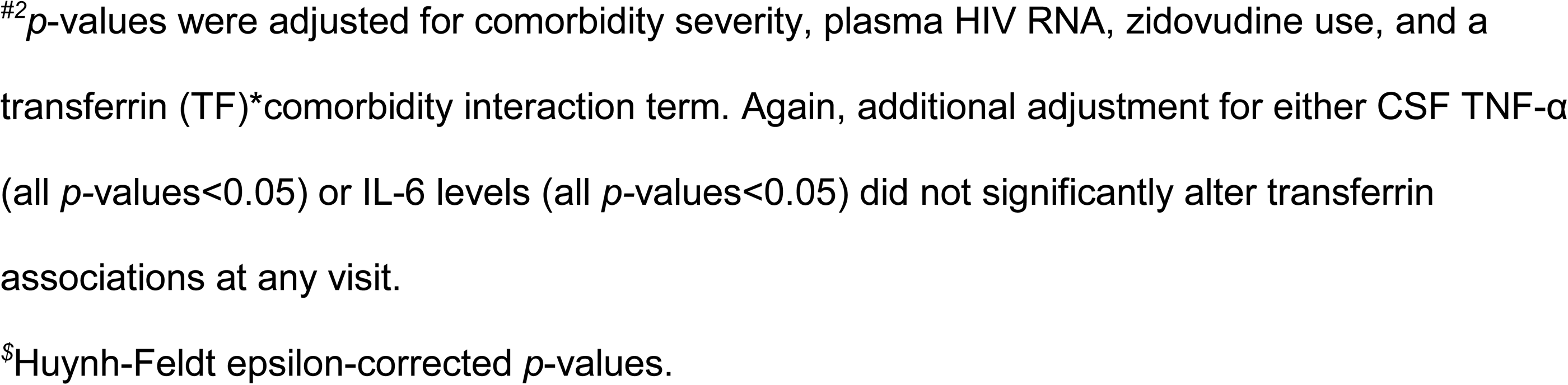
Significant associations of CSF transferrin with the GDS over time were observed primarily in adults 50 years of age and older.

| Variable | 30 months (N=41) |  | 36 months (N=37) |  | 42 months (N=34) |  |
| --- | --- | --- | --- | --- | --- | --- |
| | <sup>#2</sup>*p* | <sup>\$2</sup>*p* | <sup>#2</sup>*p* | <sup>\$2</sup>*p* | <sup>#2</sup>*p* | <sup>\$2</sup>*p* |
| Transferrin | **0.01** | **0.04** | **0.04** | 0.09 | **0.03** | **0.08** |
| Comorbidity | 0.07 | 0.10 | 0.12 | 0.17 | 0.34 | 0.34 |
| TF*Comorbidity Interaction | **<0.01** | **0.02** | **0.01** | **0.05** | **0.04** | 0.09 |
| Plasma HIV RNA | 0.17 | --- | 0.36 | --- | 0.73 | --- |
| Zidovudine use | 0.86 | --- | 0.80 | --- | 0.94 | --- |

*p*-values<0.05 (**bolded**) are statistically significant.
<sup>#2</sup>*p*-values were adjusted for comorbidity severity, plasma HIV RNA, zidovudine use, and a transferrin (TF)\*comorbidity interaction term. Again, additional adjustment for either CSF TNF-α (all *p*-values<0.05) or IL-6 levels (all *p*-values<0.05) did not significantly alter transferrin associations at any visit.
<sup>\$</sup>Huynh-Feldt epsilon-corrected *p*-values.

**S6 Table.**
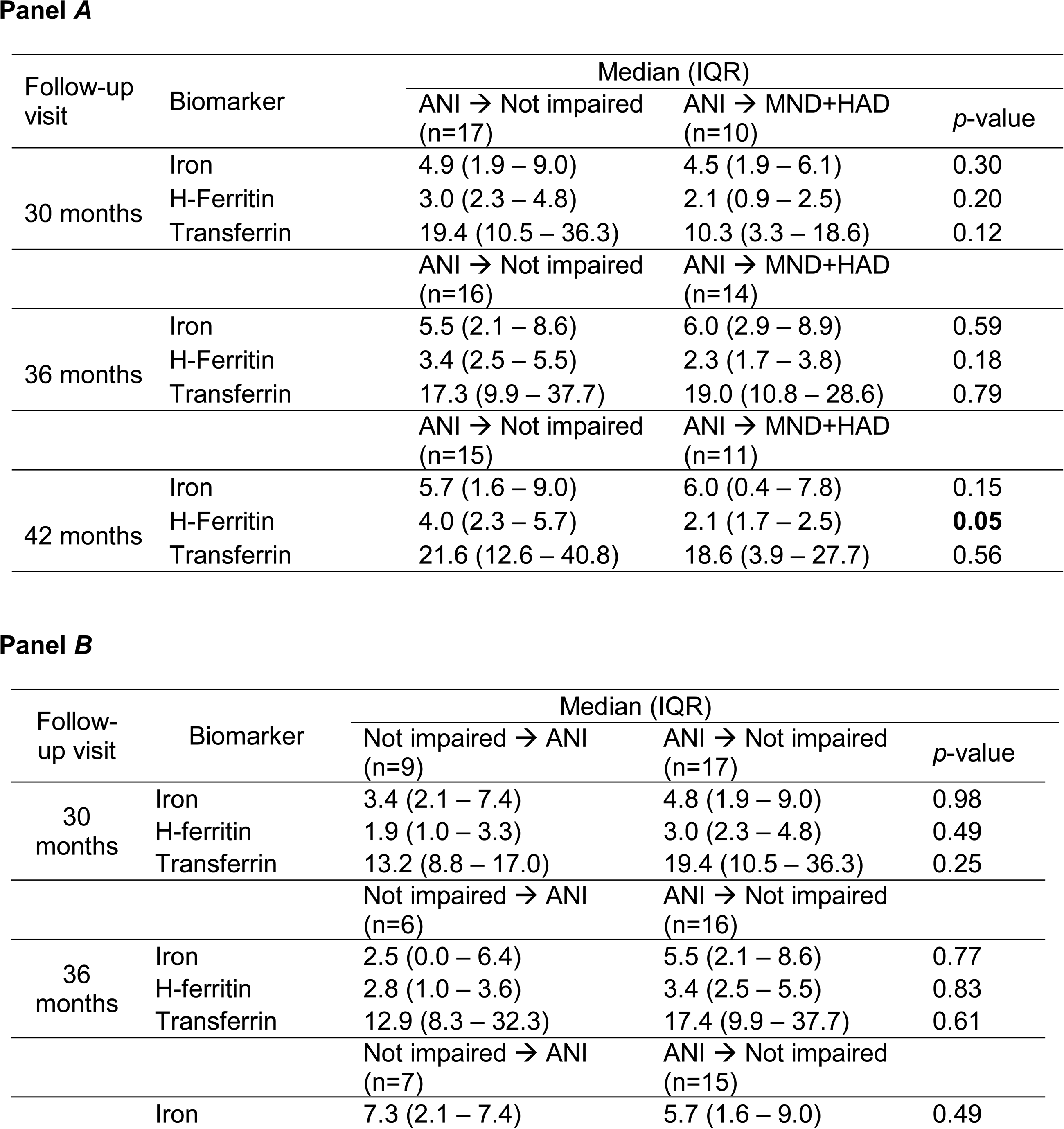
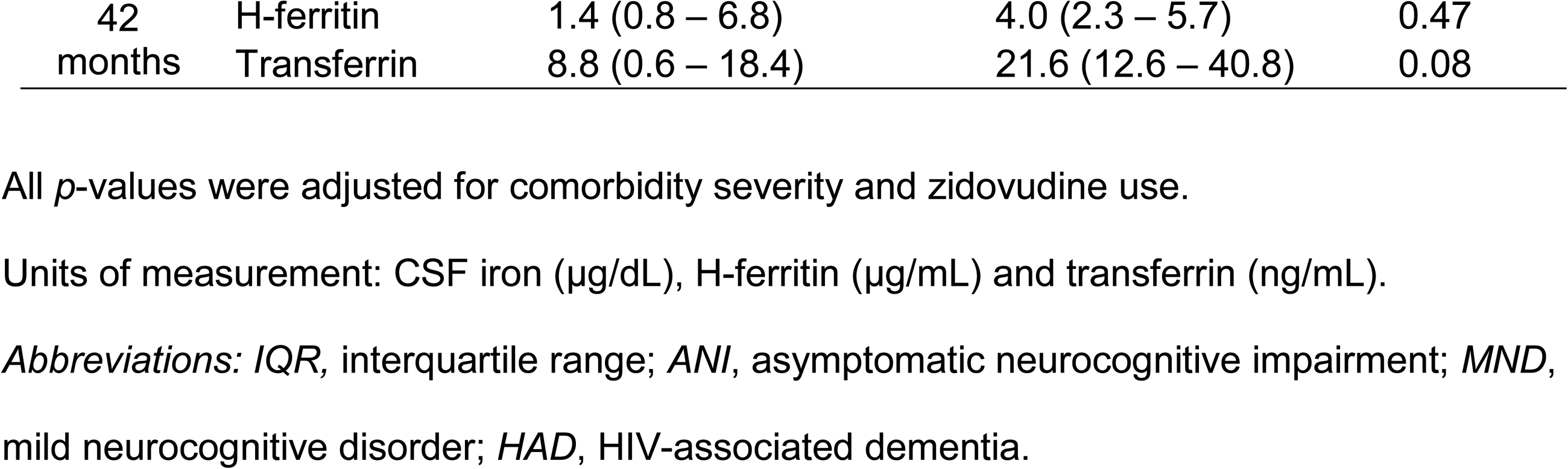
Distributions of CSF iron, H-ferritin, and transferrin levels at baseline among participants who improved from ANI at baseline to *not impaired*, as compared to individuals who converted from ANI to MND or HAD at their last follow-up visit (panel ***A***). CSF biomarker levels among participants who converted from *not impaired* at baseline to ANI, as compared to people who converted from ANI to *not impaired* at their last follow-up visit (panel ***B***).

**S7 Table.** Associations of CSF H-ferritin and transferrin with GDS differences up to and including the 42-month follow-up visit, after excluding outlier values of these biomarkers (values ≥2 SD above or below the mean) in all study participants (panel ***A***), and for H-ferritin in the subset aged <50 years (panel ***B***).

| 42-month follow-up visit |  |  |  |
| --- | --- | --- | --- |
| Model | Variable | <sup>#</sup>*p* | <sup>\$</sup>*p* |
| Model 1<sup>a</sup><br>(N=99) | H-Ferritin | **<0.01** | **<0.01** |
|  | Comorbidity | 0.45 | 0.42 |
|  | H-Ft*Comorbidity Interaction | 0.32 | 0.33 |
| Model 2<sup>b</sup><br>(N=91) | Transferrin | **<0.01** | **0.01** |
|  | Comorbidity | 0.53 | 0.49 |
|  | TF*Comorbidity Interaction | 0.11 | 0.15 |

Panel B *Study participants aged <50 years*
| 42 month follow-up visit (N=69) |  |  |
| --- | --- | --- |
| Variable | <sup>#</sup>*p* | <sup>\$</sup>*p* |
| H-Ferritin | **<0.01** | **0.01** |
| Comorbidity | 0.86 | 0.78 |
| H-Ft*Comorbidity Interaction | **0.03** | 0.07 |

*p*-values<0.05 (**bolded**) are statistically significant.
<sup>a#</sup>*p*-values were adjusted for comorbidity severity, plasma HIV RNA, and a H-ferritin (H-Ft)\*comorbidity interaction term.
<sup>b#</sup>*p*-values were adjusted for comorbidity severity, HIV RNA, zidovudine use and a transferrin (TF)\*comorbidity interaction term.
<sup>\$</sup>Huynh-Feldt epsilon-corrected *p*-values.

**S8 Table.** Associations of CSF H-ferritin (panel ***A***) and transferrin (panel ***B***) with GDS differences across three study visits in all participants, after excluding tertile 2 data from ANOVA regression models.

**Panel A**
| Variable | 30 months<br>(N=104) |  | 36 months<br>(N=86) |  | 42 months<br>(N=72) |  |
| --- | --- | --- | --- | --- | --- | --- |
| | #<sup>1</sup>*p* | \$<sup>1</sup>*p* | #<sup>1</sup>*p* | \$<sup>1</sup>*p* | #<sup>1</sup>*p* | \$<sup>1</sup>*p* |
| H-Ferritin | **<0.01** | **<0.01** | **0.03** | **0.03** | 0.12 | 0.13 |
| Comorbidity | 0.85 | 0.84 | 0.40 | 0.39 | **0.04** | **0.05** |
| H-Ft*Comorbidity<br>Interaction | 0.75 | 0.75 | 0.42 | 0.41 | 0.56 | 0.55 |

**Panel B**
| Variable | 30 months<br>(N=93) |  | 36 months<br>(N=77) |  | 42 months<br>(N=65) |  |
| --- | --- | --- | --- | --- | --- | --- |
| | #<sup>2</sup>*p* | \$<sup>2</sup>*p* | #<sup>2</sup>*p* | \$<sup>2</sup>*p* | #<sup>2</sup>*p* | \$<sup>2</sup>*p* |
| Transferrin | **<0.01** | **<0.01** | **<0.01** | **0.01** | **<0.01** | **<0.01** |
| Comorbidity | 0.07 | 0.10 | 0.15 | 0.18 | 0.21 | 0.24 |
| TF*Comorbidity<br>Interaction | **0.04** | 0.07 | **<0.01** | **0.02** | **<0.01** | **0.02** |

*p*-values<0.05 (**bolded**) are statistically significant.

**S9 Table.**
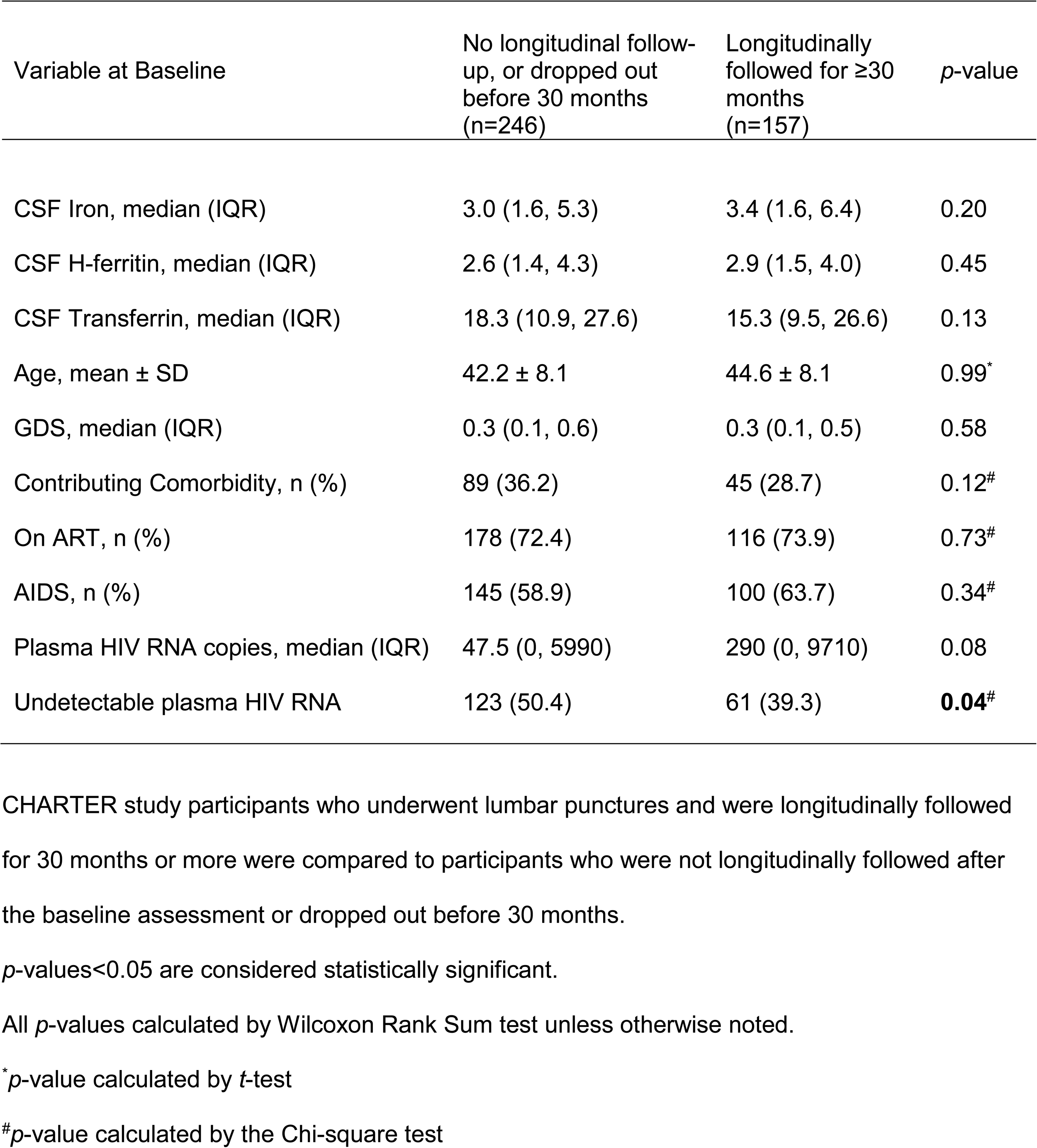
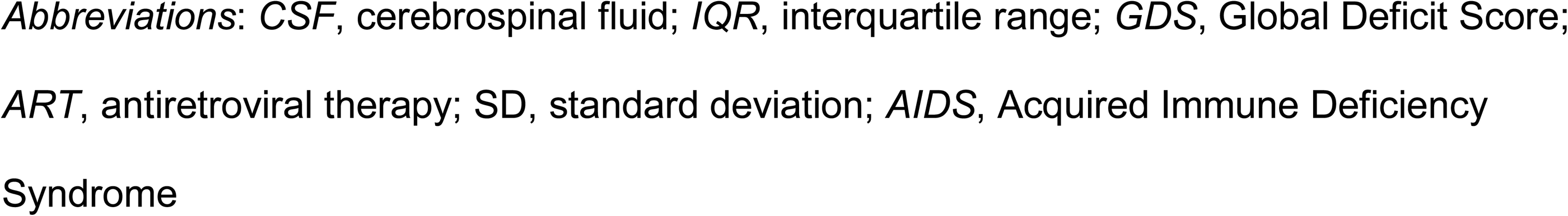
Comparison of baseline distributions of CSF iron (μg/dL), H-ferritin (μg/mL), transferrin (ng/mL), and selected demographic and HIV disease factors by participant follow-up status.

| Variable at Baseline | No longitudinal follow-up, or dropped out before 30 months (n=246) | Longitudinally followed for ≥30 months (n=157) | *p*-value |
| --- | --- | --- | --- |
| CSF Iron, median (IQR) | 3.0 (1.6, 5.3) | 3.4 (1.6, 6.4) | 0.20 |
| CSF H-ferritin, median (IQR) | 2.6 (1.4, 4.3) | 2.9 (1.5, 4.0) | 0.45 |
| CSF Transferrin, median (IQR) | 18.3 (10.9, 27.6) | 15.3 (9.5, 26.6) | 0.13 |
| Age, mean ± SD | 42.2 ± 8.1 | 44.6 ± 8.1 | 0.99* |
| GDS, median (IQR) | 0.3 (0.1, 0.6) | 0.3 (0.1, 0.5) | 0.58 |
| Contributing Comorbidity, n (%) | 89 (36.2) | 45 (28.7) | 0.12<sup>#</sup> |
| On ART, n (%) | 178 (72.4) | 116 (73.9) | 0.73<sup>#</sup> |
| AIDS, n (%) | 145 (58.9) | 100 (63.7) | 0.34<sup>#</sup> |
| Plasma HIV RNA copies, median (IQR) | 47.5 (0, 5990) | 290 (0, 9710) | 0.08 |
| Undetectable plasma HIV RNA | 123 (50.4) | 61 (39.3) | **0.04<sup>#</sup>** |

CHARTER study participants who underwent lumbar punctures and were longitudinally followed for 30 months or more were compared to participants who were not longitudinally followed after the baseline assessment or dropped out before 30 months.
*p*-values<0.05 are considered statistically significant.
All *p*-values calculated by Wilcoxon Rank Sum test unless otherwise noted.
\**p*-value calculated by *t*-test
<sup>#</sup>*p*-value calculated by the Chi-square test
*Abbreviations:* CSF, cerebrospinal fluid; IQR, interquartile range; GDS, Global Deficit Score; ART, antiretroviral therapy; SD, standard deviation; AIDS, Acquired Immune Deficiency Syndrome

**(S1-S9 Tables DOCX)**

---

**STROBE-ME Table**

**(DOCX)**

**STROBE-ME: The STrengthening in Reporting OBservational studies in Epidemiology – Molecular Epidemiology (STROBE-ME) Reporting Recommendations: Extended from STROBE statement**

Reporting Recommendations: Extended from STROBE statement
| Item | Item number | STROBE Guidelines | Extension for Molecular Epidemiology Studies (STROBE-ME) | Page/Line No | Relevant text from manuscript |
| --- | --- | --- | --- | --- | --- |
| **Title and abstract** | 1 | (a) Indicate the design of the study with a commonly used term in the title or the abstract | ME-1 State the use of specific biomarker(s) in the title and/or in the abstract if they contribute substantially to the findings | Lines 31-36, 107-109 | See Title wording, and Abstract |
|  |  | (b) Provide in the abstract an informative and balanced summary of what was done and what was found |  | Lines 34-54 | See Abstract |
| **Introduction** |  |  |  |  |  |
| Background rationale | 2 | Explain the scientific background and rationale for the investigation being reported | ME-2 Explain in the scientific background of the paper how/why the specific biomarker(s) have been chosen, potentially among many others (e.g. others are studied but reported elsewhere or not studied at all) | Lines 27-28, 31-34, 75-114 (especially 90-105) | Abstract and Introduction |
| Objectives | 3 | State specific objectives, including any pre-specified hypotheses | ME-3 A priori hypothesis: if one or more biomarkers are used as proxy measures, state the *a priori* hypothesis on the expected values of the biomarker(s) | Lines 31-34, 107-111 | Abstract and Introduction |
| **Methods** |  |  |  |  |  |
| Study design | 4 | Present key elements of study design early in the paper | ME-4 Describe the special study designs for molecular epidemiology (in particular, nested case/control and case/cohort) and how they were implemented | Figure 4<br>Lines 141-144, 167-169, 216-219, 231-238, 407-413, 417-433, 447-511 | Methods and Results |
| Biological sample collection |  |  | ME-4.1 Report on the setting of the biological sample collection; amount of sample; nature of collecting procedures; participant conditions; time between sample collection and relevant clinical or physiological endpoints | Lines 410-413, 440-444 | All samples were collected between 8 am and 2:30 pm, depending on study requirements, and other study visits for participants that day. Volume of CSF drawn was 16 mL, always within 1 hour of blood collection. |
| Biological sample storage |  |  | ME-4.2 Describe sample processing (centrifugation, timing, additives, etc.) |  | Samples were refrigerated immediately and processed within a 1-hour window of collection. CSF was centrifuged at 250xg for 15 minutes. CSF supernatant was transferred to cryovials in 1 or 0.5 mL aliquots. CSF supernatant was stored at -80°C. No additives were used. |
| Biological sample processing |  |  | ME-4.3 Describe sample storage until biomarker analysis (storage, thawing, manipulation, etc.) |  | No additives or manipulation. No freeze-thaws until assays were performed. Specimens were stored at -80°C until sent out (frozen, on dry ice) for laboratory assays. |
| Biomarker biochemical characteristics |  |  | ME-4.4 Report the half-life of the biomarker and chemical and physical characteristics (e.g. solubility) | Lines 95-96 | See Methods: Iron, transferrin, and ferritin are longstanding assays/biomarkers of known physicochemical characteristics and *ex vivo* stability in human specimens (References 1-4, *below*. PMIDs:18991401; 29228047; 31503347; 23627617) |
| Setting | 5 | Describe the setting, locations and relevant dates, including periods of recruitment, exposure, follow-up and data collection |  | Lines 407-408, 417-433, 466-468 | Methods and Results |
| Participants | 6 | (a) Cohort study – Give the eligibility criteria and the sources and methods of selection of participants. Describe methods of follow-up Case-control study – Give the eligibility criteria and the sources and methods of case ascertainment and control selection. Give the rationale for the choice of cases and controls<br>Cross-sectional study – Give the eligibility criteria and the sources and methods of selection of participants | ME-6 Report any habit, clinical condition, physiological factor or working or living condition that might affect the characteristics or concentrations of the biomarker | Cohort study: Lines 141-142, 167-169, 407-414, 417-419, 426-432; Lines | Methods and Results<br>Only baseline analysis was cross-sectional; all other analyses were longitudinal/prospective. |
|  |  | (b) Cohort study – For matched studies, give matching criteria and number of exposed and unexposed |  | N/A | ---- |
|  |  | Case-control study – For matched studies, give matching criteria and the number of controls per case |  |  |  |
| Variables | 7 | Clearly define all outcomes, exposures, predictors, potential confounders and effect modifiers. Give diagnostic criteria, if applicable |  | Lines 416-433, 435-440, 446-511 | Methods |
| Data source/<br>measurement | 8 | For each variable of interest, give sources of data and details of methods of assessment (measurement). Describe comparability of assessment methods if there is more than one group | ME-8 Laboratory methods: report type of assay used, detection limit, quantity of biological sample used, outliers, timing in the assay procedures (when applicable) and calibration procedures or any standard used | Lines 407-488; 497-498; Tables S7 A and B | See Methods (Assessment of HAND/NCI, Quantification of inflammation, Statistical Analysis); Assay characteristics are previously published and referenced (Reference 5, *below*; PMID:28427421) |
| Bias | 9 | Describe any efforts to address potential sources of bias |  | Lines 443-444, 502-505<br>Table S9<br>Lines 247-259 & 492-505 | CSF assays blinded to outcome;<br>Table S9 shows results of comparison of CSF measures and clinical variables in evaluable participants at 30 months vs. people who did not have follow-up data at 30 months)<br>Numerous methods to assess robustness of findings |
| Study size | 10 | Explain how the study size was arrived at |  | Figure 4<br>Lines 410-413, 141-142, 167-169 | See Methods and Results |
| Quantitative variables | 11 | Explain how quantitative variables were handled in the analyses. If applicable, describe which groupings were chosen and why |  | Lines 446-515<br>Pages 20-23 | See detailed Statistical Methods |
| Statistical methods | 12 | (a) Describe all statistical methods, including those used to control for confounding | ME-12 Describe how biomarkers were introduced into statistical models | Lines 446-515 | See detailed statistical section Biomarkers were introduced as “continuous” variables in logistic regression and “tertiles” in ANOVA analyses. |
|  |  | (b) Describe any methods used to examine subgroups and interactions |  | Figure 4 | See detailed statistical section |
|  |  | (c) Explain how missing data were addressed |  | Table S9 | See detailed statistical section There were no appreciable missing data, as noted in Table 1 legend. |
|  |  | (d) Cohort study – If applicable, explain how loss to follow-up was addressed<br>Case-control study – If applicable, explain how matching of cases and controls was addressed<br>Cross-sectional study – If applicable, describe analytical methods taking account of sampling strategy |  | Line 446-515 and | See detailed statistical section; also Table S9 |
|  |  | (e) Describe any sensitivity analyses |  | Lines 231-245, 247-259, 377-380, 385-392, 485-494<br>Tables S2, S6, S7 and S8 | The Huynh-Feldt correction was applied to longitudinal analyses; Analyses repeated with CSF biomarker outliers excluded; Analyses of neurocognitive function and changes therein were performed in several different ways with consistent results |
| Validity/<br>reliability of<br>measurement<br>and internal/<br>external<br>validation |  |  | ME-12.1 Report on the validity and reliability of measurement of the biomarker(s) coming from the literature and any internal or external validation used in the study | Lines 385-388 (robustness of findings) | See Discussion;<br>See also References 1-5, *below*. |
| **Results** |  |  |  |  |  |
| Participants | 13 | (a) Report the numbers of individuals at each stage of the study – e.g. numbers potentially eligible, examined for eligibility, confirmed eligible, included in the study, completing follow-up and analyzed | ME-13 Give reason for loss of biological samples at each stage | Figure 4<br>Lines 119-127, 141-142, 166-169 | No loss of biological samples at any stage |
|  |  | (b) Give reasons for nonparticipation at each stage |  | Table S9 | See Supplemental material |
|  |  | (c) Consider use of a flow diagram |  | Figure 4<br>Lines 167-169 | N/A |
| Descriptive data | 14 | (a) Give characteristics of study participants (e.g. demographic, clinical and social) and information on exposures and potential confounders |  | Table 1<br>Lines 119-127, 130-138, 141-144 | See Results |
|  |  | (b) Indicate the number of participants with missing data for each variable of interest |  | Tables 1 and 2 legends | See results |
|  |  | (c) Cohort study – Summarize follow-up time (e.g. average and total amount) |  | Lines 141-142, 167-169 | See Results |
| Distribution of biomarker measurement |  |  | ME-14.1 Give the distribution of the biomarker measurement (including mean, median, range and variance) | Table 2<br>Lines 130-133 | See Results |
| Outcome data | 15 | Cohort study – Report numbers of outcome events or summary measures over time<br>Case-control study – Report numbers in each exposure category or summary measures of exposure<br>Cross-sectional study – Report numbers of outcome events or summary measures |  | Lines 119-125, 133-138, 141-143, 167-169 | See Results |
| Main results | 16 | (a) Give unadjusted estimates and, if applicable, confounder-adjusted estimates and their precision (e.g. 95% confidence interval). Make clear which confounders were adjusted for and why they were included |  | Tables 3-5<br>S2-S8<br>Lines 133-138, 141-163 | All Statistical section, Results, Main and Supplementary tables |
|  |  | (b) Report category boundaries when continuous variables were categorized |  | Lines 130-133 | See Results |
|  |  | (c) If relevant, consider translating estimates of relative risk into absolute risk for a meaningful time period |  | N/A | --- |
| Other analyses | 17 | Report other analyses done – e.g. analyses of subgroups and interactions and sensitivity analyses |  | Figures 1 and 4;<br>Lines 151-155, 157-163, 179-191, 200-205, 207-213, 231-245, 248-259 | See main Results and Supplemental Information |
| **Discussion** |  |  |  |  |  |
| Key results | 18 | Summarize key results with reference to study objectives |  | Lines 107-114, 284-289, 394-401 | See (Abstract), Introduction, and Discussion |
| Limitations | 19 | Discuss limitations of the study, taking into account sources of potential bias or imprecision. Discuss both direction and magnitude of any potential bias | ME-19 Describe main limitations in laboratory procedures | Lines 373-392, 399-401 | Design limitations discussed in Discussion section |
| Interpretation | 20 | Give a cautious overall interpretation of results considering objectives, limitations, multiplicity of analyses, results from similar studies and other relevant evidence | ME-20 Give an interpretation of results in terms of a priori biological plausibility | Lines 271-283, 346-349 | See Discussion, including differences from animal models |
| Generalizability | 21 | Discuss the generalizability (external validity) of the study results |  | Lines 396-401 | Discussion |
| **Other information** |  |  |  |  |  |
| Funding | 22 | Give the source of funding and the role of the funders for the present study and, if applicable, for the original study on which the present article is based |  | Lines 8-11 | See Funding Disclosures |
| Ethics |  |  | ME-22.1 Describe informed consent and approval from ethical committee(s). Specify whether samples were anonymous, anonymized or identifiable | Lines 408-410, 413-414<br>All the samples were anonymized | See Methods |

## Notes

**Funding Disclosure:** This work was supported by National Institutes of Health grant 1R01 MH095621 (to AK and TH). The CNS HIV Anti-Retroviral Therapy Effects Research (CHARTER) Study was also supported by National Institutes of Health awards NIH R01 MH107345 (to SL and RH).

### Competing Interest Statement

Authors Kaur, Bush, Letendre, Franklin, Ellis, Hulgan, Heaton, Samuels, and Kallianpur have no potential conflicts of interest to disclose. Author S. Patton is a paid consultant for SideroBiosciences, Inc., and J.R. Connor, a longstanding collaborator, is Chairman of the Board and co-founder of SideroBiosciences, which has a product in clinical trials for treating iron deficiency.

### Funding Statement

This work was supported by National Institutes of Health grant 1R01 MH095621 (to AK and TH). The CNS HIV Anti-Retroviral Therapy Effects Research (CHARTER) Study was also supported by National Institutes of Health awards NIH R01 MH107345 (to SL and RH). The authors and their institutions received no payment or services from a third party for any aspect of the submitted work.

### Author Declarations

Cleveland Clinic Institutional Review Board approval was given for this analysis, which was considered exempt due to using only completely de-identified data. The CHARTER Study on which the analysis is based has been continuously approved by the IRBs of all six participating institutions. Further details will be provided as needed.

